# Immune checkpoint blockade reshapes drug-associated toxicity: a pharmacovigilance atlas of drug-ICI interactions

**DOI:** 10.64898/2026.08.31.26361880

**Authors:** Eric Milan Mukherjee, Amir Asiaee, Dodie Park, Matthew S. Krantz, Cosby A. Stone, Michelle Martin-Pozo, Elizabeth Phillips

## Abstract

**Background:** Immune checkpoint inhibitors (ICIs) are usually treated as direct culprits in immune-related adverse events, but checkpoint blockade may also reset tolerance to other medications. We tested whether ICI exposure reshapes the organization, drug specificity and timing of reported treatment toxicity.

**Methods:** We analyzed 13,701,106 deduplicated FDA Adverse Event Reporting System reports from 2016 through 2025. Cancer-restricted reporting associations and cross-organ community detection characterized the ICI-associated toxicity landscape. Adjusted logistic models tested primary-suspect drug x ICI interactions for Stevens-Johnson syndrome/toxic epidermal necrolysis (SJS/TEN), drug reaction with eosinophilia and systemic symptoms (DRESS), acute generalized exanthematous pustulosis, interstitial nephritis, drug-induced liver injury, anaphylaxis and vomiting. Accelerated failure-time models evaluated documented onset according to ICI exposure and checkpoint pathway.

**Results:** Among 2,365,278 cancer-associated reports, 256,940 contained an ICI. Of 3001 eligible Preferred Terms, 2091 differed between ICI-containing and non-ICI reports at false discovery rate (FDR) <0.05, and four cross-organ toxicity communities emerged. Drug-phenotype associations were reweighted: 71 of 147 eligible pairs had FDR-significant interactions, including 56 amplifications and 15 attenuations. Signals included marked moxifloxacin-SJS/TEN amplification (interaction OR 100.51, 95% CI 38.58 to 261.84), a submultiplicative enfortumab vedotin-SJS/TEN interaction (0.17, 0.13 to 0.23) and omeprazole-interstitial nephritis amplification (10.33, 7.60 to 14.04). Among 64,593 reports with documented onset of 1-365 days, ICI exposure was associated with longer adjusted onset for five of seven phenotypes (time ratios 1.37-1.58); 10 of 19 estimable checkpoint-phenotype coefficients remained FDR significant.

**Conclusions:** Checkpoint blockade was associated not simply with additional toxicity, but with a change in treatment context: drug-phenotype associations shifted in both directions, adverse events formed cross-organ structure and documented onset varied by checkpoint pathway. These findings support ICIs as modifiers of drug toxicity and identify specific signals for longitudinal and mechanistic validation.

**KEY MESSAGES:** *WHAT IS ALREADY KNOWN ON THIS TOPIC:*

- Experimental PD-1/PD-L1 blockade can lower the threshold for drug-specific T-cell priming, and hypersensitivity to previously tolerated medications has emerged after ICI exposure. Whether this immune rewiring systematically changes how other drugs produce toxicity across phenotypes is unknown.

*WHAT THIS STUDY ADDS:*

- Across 13,701,106 deduplicated FAERS reports, 71 of 147 eligible primary-suspect drug-phenotype pairs had FDR-significant drug x ICI interactions, with amplification and attenuation differing sharply by phenotype. Cross-organ organization and checkpoint-dependent timing provided complementary signatures of an altered treatment state.

*HOW THIS STUDY MIGHT AFFECT RESEARCH, PRACTICE OR POLICY:*

- The findings support a two-hit treatment-state model: checkpoint blockade may alter immune permissiveness, while a second medication helps determine whether and how toxicity is expressed. Individual signals require longitudinal and mechanistic validation and should not be used to estimate incidence or justify drug avoidance from spontaneous-reporting data alone.

## Introduction

Immune checkpoint inhibitors (ICIs) transform cancer therapy by releasing inhibitory signals that normally constrain T-cell activation.^1–2^ The same biology disrupts immune tolerance and produces immune-related adverse events (irAEs).^3–4^ Yet checkpoint therapy is rarely pharmacologically isolated. Patients are simultaneously exposed to cytotoxic and targeted anticancer agents, antimicrobials, acid suppressants, analgesics, anticoagulants and medications introduced as treatment evolves. Most safety frameworks place the ICI at the center and treat these other drugs as background exposures. But if checkpoint blockade alters immune tolerance, it may do more than add another potential culprit: it may change the rules under which the rest of the regimen is tolerated.

The two-hit hypothesis has direct experimental support. PD-1/PD-L1 blockade lowers the threshold for priming naive T cells against drug-associated antigens and increases drug hypersensitivity.^5–6^ Clinically, hypersensitivity to previously tolerated medications has emerged after checkpoint blockade, including reactions to iodinated contrast and targeted anticancer therapies.^7–10^ In our previous analysis of SJS/TEN, culprit drugs interacted with ICI exposure and latency differed across checkpoint pathways.^11^ Together, these observations suggest a first hit that changes immune permissiveness and a second exposure that helps determine whether, where and how toxicity is expressed. Crucially, this model does not predict uniform amplification. The effect should depend on the drug, phenotype and checkpoint pathway, while drugs dominated by direct or nonimmune toxicity may behave differently from classical T-cell-mediated culprits.

Several checkpoint effects could widen this permissive state. Checkpoint inhibition expands or reinvigorates T-cell clones, including populations recognizing antigens shared by tumor and normal tissue; perturbs circulating B-cell compartments; and produces tissue-specific inflammatory programs against a background of systemic immune activation.^12–15^ Intestinal microbial states are also associated with ICI response and toxicity, providing another route through which antibiotics and other concomitant exposures could modify immune behavior.^16–18^ Multiorgan irAEs and recognizable overlap syndromes provide additional evidence that checkpoint blockade perturbs immune context beyond a single organ.^19–20^ These observations make a purely additive model, in which every drug carries an unchanged toxicity profile regardless of checkpoint exposure, increasingly incomplete.

Testing that model requires interaction analysis, not simply a longer catalogue of irAEs. A two-hit process should leave several complementary footprints: the overall distribution of toxicity should shift with ICI exposure; the association between an individual drug and a specific phenotype should change in the presence of an ICI; and the timing of toxicity may vary with ICI exposure or checkpoint pathway. Cross-organ organization provides a separate view of the altered state, but no single signature is sufficient because each can reflect reporting behavior, case mix or regimen composition. We therefore analyzed 13.7 million deduplicated FAERS reports from 2016 through 2025 to construct a pharmacovigilance atlas of drug-ICI interactions, moving from the broader toxicity landscape to drug-specific interactions and finally to checkpoint-dependent temporality.

## Methods

### Study design, data source and preprocessing

We performed a cross-sectional study of FAERS reports from 2016 through 2025, using the report as the unit of analysis. Data were imported and deduplicated using the faers R/Bioconductor package.^21^ Drug labels were standardized to canonical ingredients; ICI exposure was assigned with a curated checkpoint-target dictionary and was considered present in any reported drug role unless otherwise specified.

Adverse events used Medical Dictionary for Regulatory Activities (MedDRA) v29.0 Preferred Terms (PTs) and Higher Level Terms (HLTs). Cancer-associated reports were identified from manually reviewed malignant-indication and cancer-specific drug-proxy dictionaries. Detailed preprocessing, drug adjudication, cancer assignment, phenotype definitions, age imputation, and software specifications are provided in **eMethods**. Because FAERS contains publicly available, deidentified reports, institutional review board approval was not required.

The analyses were aligned to complementary predictions of an ICI-altered treatment state. The reporting-association analysis asked whether the overall adverse-event distribution shifted with ICI exposure; the network analysis asked whether events co-reported with ICIs formed reproducible cross-organ structure; and the interaction models asked whether the association between a primary-suspect drug and a focal phenotype differed according to ICI exposure. Reporting ORs and interaction ORs are measures of relative reporting, not estimates of incidence.^22–23^ Temporal models supplied an independent dimension by testing documented onset rather than reporting frequency.

### ICI-associated adverse-event landscape and cross-organ communities

The primary PT association analysis was restricted to cancer-associated reports and compared any-role ICI exposure with no ICI exposure. PTs reported in at least 50 cancer-associated reports overall and at least 10 ICI-containing reports were eligible. Reporting ORs and 95% CIs were calculated from 2 × 2 tables; likelihood-ratio P values and Benjamini-Hochberg adjustment used 256-bit arbitrary-precision log-space arithmetic (**eTable 5; eFigure 3**).

Within all ICI-containing reports, PTs were collapsed to HLTs and HLTs reported in at least 50 ICI-containing reports were retained. Network edges required prespecified support, lift, phi, normalized pointwise mutual information, and equal-report-weight sensitivity criteria; contextual nonbiological

MedDRA system organ classes were excluded. Communities were identified using the Leiden algorithm and retained as cross-organ when they met prespecified size, organ-system diversity, and cross-system connectivity criteria. Full network definitions and thresholds are provided in eMethods (**eTables 6-9; eFigure 4**). The network was constructed only within ICI-containing reports so that communities represented the internal organization of reported ICI toxicity rather than separation between exposed and unexposed reports. Multiple edge criteria were required because raw co-report counts favor common events, whereas lift and normalized pointwise mutual information can overemphasize rare pairs. The equal-report-weight sensitivity analysis reduced the influence of reports containing many adverse-event terms.

### Drug x ICI interaction analysis

We evaluated 7 focal phenotypes: Stevens-Johnson syndrome/toxic epidermal necrolysis (SJS/TEN), drug reaction with eosinophilia and systemic symptoms (DRESS), acute generalized exanthematous pustulosis (AGEP), interstitial nephritis, drug-induced liver injury (DILI), anaphylaxis, and vomiting. Anaphylaxis was included as an immune-mediated but non-T-cell-dominant comparator and vomiting as a nonimmune control. For each phenotype, non-ICI primary-suspect ingredients were eligible if at least 15 phenotype reports contained the drug and at least 5 also contained an ICI. The primary analysis used adjusted logistic regression with primary-suspect drug exposure, any-role ICI exposure, and their interaction, adjusting for age, original age missingness, sex, geographic region, calendar year, number of reported drugs, and cancer status. The exponentiated interaction coefficient estimates how the drug-phenotype reporting association differs between ICI-containing and non-ICI reports. FDR adjustment was performed separately within phenotype. Robustness was assessed using complete-case age models and 5000 repeated 80% subsamples, summarized by directional recovery and coefficient stability; detailed model and subsampling specifications are provided in eMethods (**eTables 10-11**; **eFigure 5**; **Figure 5**).

The focal phenotypes were selected before modeling to span severe T-cell-mediated reactions, organ-specific immune injury, a predominantly immediate hypersensitivity comparator and a common nonimmune symptom. Eligibility thresholds were applied before model fitting to exclude drug-phenotype cells too sparse for interpretable interaction estimates. The primary-suspect drug was treated as the putative phenotype-specific exposure, whereas ICI exposure was role agnostic because reporters may assign an ICI as primary suspect, secondary suspect, concomitant or interacting depending on their causal interpretation.

### Temporal analyses

Temporal analyses included phenotype-positive reports with a complete event date and at least 1 exact-date primary-suspect treatment start. The closest positive primary-suspect treatment start preceding the event was selected, and intervals of 1 to 365 days were retained for the primary analysis. Phenotype-specific accelerated failure-time models compared reports with and without any-role ICI exposure and adjusted for age, age missingness, sex, region, year, number of drugs, and cancer status; the best-fitting parametric distribution was selected by Akaike information criterion. A sensitivity analysis removed the 365-day upper bound (**eTable 12; eFigure 6**). Checkpoint-pathway models simultaneously included supported PD-1, PD-L1, CTLA-4, LAG-3, TIGIT, and TIM-3 indicators, with global Benjamini-Hochberg adjustment across estimable coefficients (eTable 13). Full temporal modeling details are provided in **eMethods**.

## Results

### Checkpoint blockade redistributes adverse-event reporting

Among 13,701,106 deduplicated reports, 2,365,278 were classified as cancer associated, including 256,940 containing an ICI (**eFigures 1-2; eTables 1-4**). Median age among cancer reports with observed age was 66 years (IQR, 56-75 years). PD-1 agents were most frequently represented, followed by PD-L1 and CTLA-4 agents; pembrolizumab and nivolumab were the most frequently reported individual ICIs. The annual number of ICI-agent report records increased from 10,717 in 2016 to 49,046 in 2025, while LAG-3, TIGIT, and TIM-3 exposure remained sparse. The cancer-restricted analysis included 3001 eligible PTs, of which 2091 differed between ICI-containing and non-ICI reports at FDR < .05: 1032 had higher reporting odds with ICI exposure and 1059 had lower reporting odds (**Figure 1**; **eTable 5**). A full-FAERS sensitivity analysis showed a similar overall distribution (**eFigure 3**). Higher-reporting PTs included hypothyroidism (OR, 13.84; 95% CI, 13.27-14.44), colitis (OR, 6.97; 95% CI, 6.72-7.24), pneumonitis (OR, 6.11; 95% CI, 5.91-6.33), myocarditis (OR, 22.58; 95% CI, 21.00-24.29), and tubulointerstitial nephritis (OR, 13.60; 95% CI, 12.55-14.73). Lower-reporting PTs included hair disorder (OR, 0.02; 95% CI, 0.01-0.03), madarosis (OR, 0.02; 95% CI, 0.01-0.03), blood count abnormal (OR, 0.05; 95% CI, 0.04-0.07), and injection-site pain (OR, 0.06; 95% CI, 0.04-0.07). These estimates describe relative reporting and do not indicate protection.

**Figure 1.**
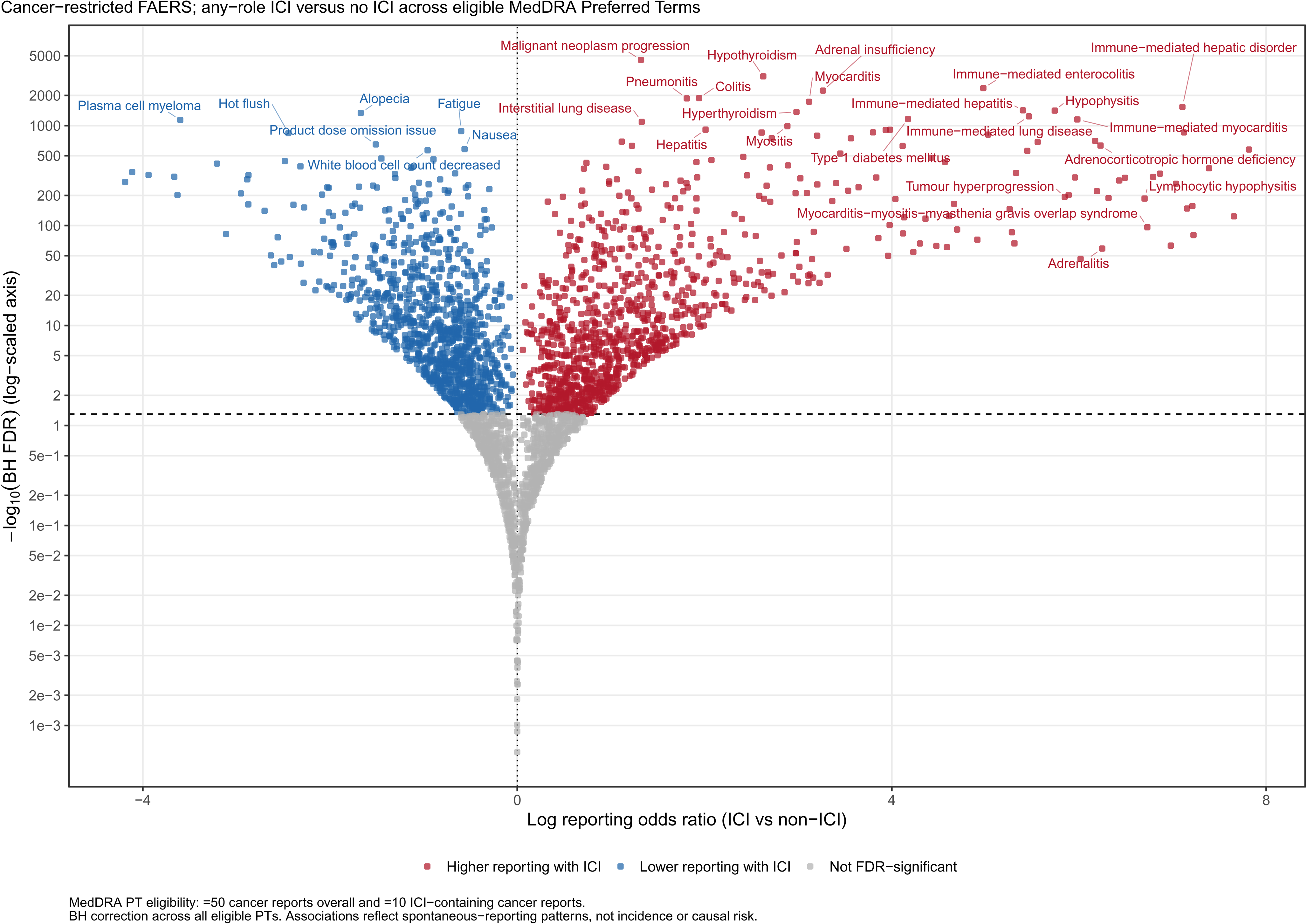
Adverse-Event Reporting Patterns Associated With Immune Checkpoint Inhibitor Exposure. Cancer-restricted Preferred-Term (PT) association analysis comparing 256,940 ICI-containing reports with 2,108,338 reports without ICI exposure among 2,365,278 cancer-associated FAERS reports. Each point represents an eligible MedDRA Preferred Term reported in at least 50 cancer-associated reports overall and at least 10 ICI-containing reports. The x-axis represents the log reporting OR for any-role ICI exposure; the y-axis represents -log10 Benjamini-Hochberg FDR displayed on a logarithmic scale. FDR correction was performed across all 3001 eligible PTs using arbitrary-precision log-space calculations. Reporting associations do not estimate incidence or causal risk.

**Figure 2.**
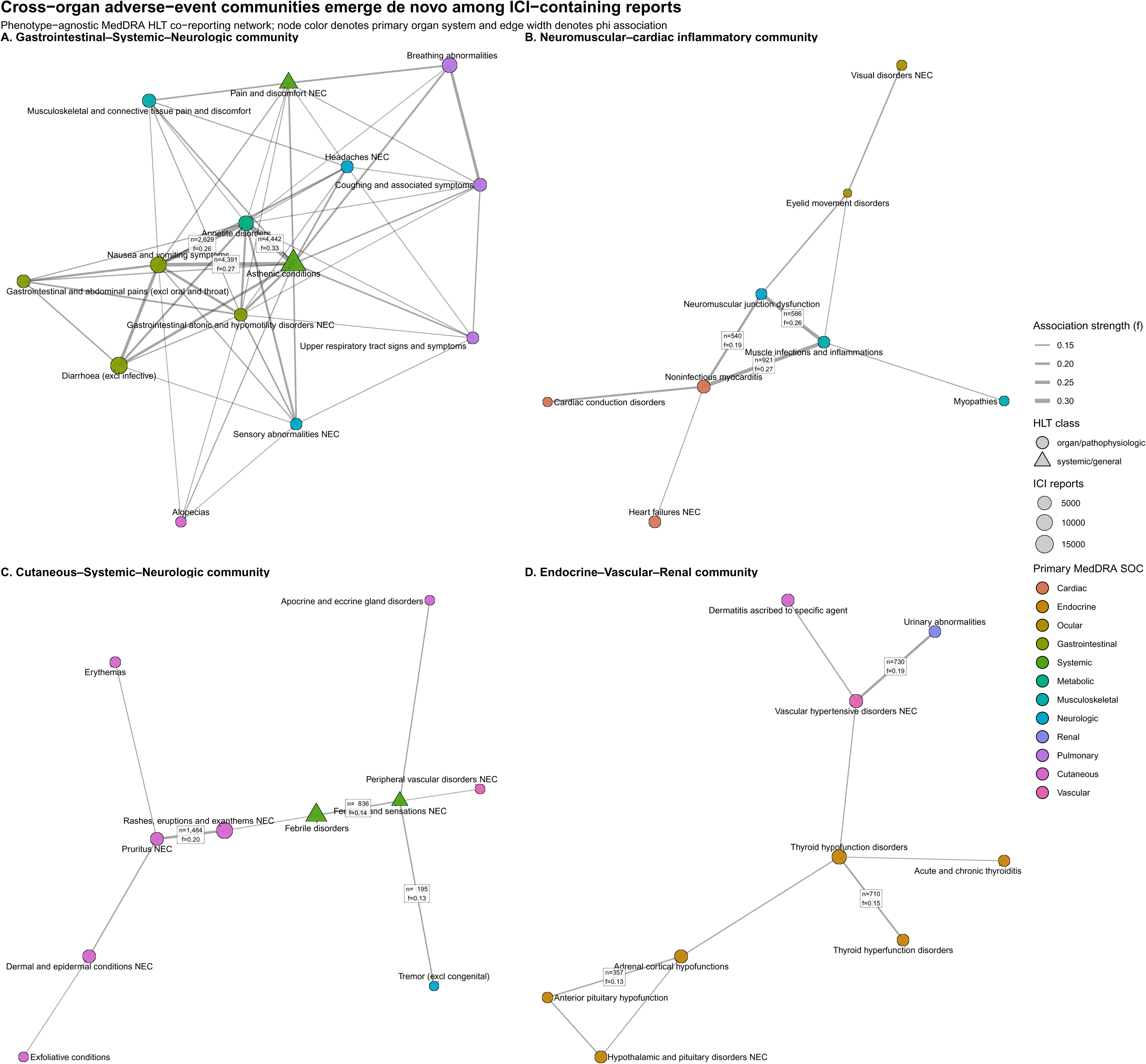
Cross-Organ Organization of Adverse Events Within ICI-Containing Reports. Four HLT communities meeting prespecified cross-organ criteria among 256,940 ICI-containing reports. Panels show the gastrointestinal-systemic-neurologic, neuromuscular-cardiac inflammatory, cutaneous-systemic-neurologic, and endocrine-vascular-renal communities. Nodes represent MedDRA Higher-level Terms (HLTs); node size reflects ICI report support, color denotes primary system organ class, and shape distinguishes organ/pathophysiologic HLTs from systemic/general HLTs. Edges represent co-reporting relationships meeting prespecified thresholds for support, lift, phi, normalized pointwise mutual information, and equal-report-weight sensitivity; edge width reflects phi.

### Four cross-organ communities emerge within ICI-containing reports

Within the 256,940 ICI-containing reports, 4 communities met prespecified cross-organ criteria (**Figure 2**; **eTables 6-9; eFigure 4**). The largest gastrointestinal-systemic-neurologic community contained 24 HLTs spanning 7 system organ classes; 55 of 70 internal edges (78.6%) crossed system organ classes. Its highest-support HLTs included asthenic conditions, diarrhea, nausea and vomiting symptoms, colitis, appetite disorders, and breathing abnormalities. A second neuromuscular-cardiac inflammatory community contained 8 HLTs spanning 4 system organ classes, with 5 of 9 internal edges crossing system organ classes. It included noninfectious myocarditis, muscle inflammation, myopathies, neuromuscular junction dysfunction, cardiac conduction disorders, heart failure, visual disorders, and eyelid movement disorders.

The cutaneous-systemic-neurologic community contained 10 HLTs from 4 system organ classes; 4 of 9 internal edges crossed system organ classes. It included rashes, pruritus, erythema, exfoliative and other dermal conditions, fever, abnormal sensations, tremor, and peripheral vascular disorders. The endocrine-vascular-renal community contained 9 HLTs from 4 system organ classes; 3 of 9 internal edges were cross-organ. Thyroid, adrenal, and pituitary disorders clustered with hypertension, urinary abnormalities, and dermatitis attributed to a specific agent.

### Drug x ICI interactions differ sharply by phenotype

Across 147 eligible drug-phenotype pairs, 71 had FDR-significant drug x ICI interactions: 56 amplifications and 15 attenuations (**Table 1**; **Figure 3**; **eTable 10**). In SJS/TEN, 15 of 27 eligible pairs were significant, including 13 amplified and 2 attenuated interactions. Moxifloxacin showed the largest amplified estimate (interaction OR, 100.51; 95% CI, 38.58-261.84; 9 jointly exposed cases), whereas enfortumab vedotin showed a submultiplicative interaction (interaction OR, 0.17; 95% CI, 0.13-0.23; 72 jointly exposed cases). In DRESS, 8 of 10 eligible pairs were significant and all 8 were amplified. Apixaban had the largest estimate (interaction OR, 117.82; 95% CI, 44.89-309.20; 5 jointly exposed cases); additional signals included lansoprazole (15.87; 95% CI, 8.00-31.48; 10 jointly exposed cases) and esomeprazole (17.08; 95% CI, 7.71-37.85; 7 jointly exposed cases). Interstitial nephritis had 18 significant interactions among 27 eligible pairs, including 11 amplified and 7 attenuated interactions. Omeprazole was amplified (10.33; 95% CI, 7.60-14.04; 89 jointly exposed cases), whereas pemetrexed was attenuated (0.44; 95% CI, 0.29-0.68; 128 jointly exposed cases).

**Table 1.** Phenotype-Specific Drug x ICI Interactions and Temporal Associations.

| Phenotype | Significant / eligible pairs<br>(amplified / attenuated) | Representative interaction ORs | Adjusted time ratio<br>(95% CI) |
| --- | --- | --- | --- |
| SJS/TEN | 15 / 27 (13 / 2) | Moxifloxacin, 100.51; enfortumab vedotin, 0.17 | 1.37 (1.21-1.54) |
| DRESS | 8 / 10 (8 / 0) | Apixaban, 117.82; esomeprazole, 17.08 | 1.43 (1.21-1.69) |
| AGEP | 1 / 1 (1 / 0) | Carboplatin, 2.39 | 0.99 (0.67-1.45) |
| Interstitial nephritis | 18 / 27 (11 / 7) | Omeprazole, 10.33; pemetrexed, 0.44 | 1.51 (1.19-1.90) |
| DILI | 10 / 18 (10 / 0) | Venlafaxine, 127.31; mycophenolate mofetil, 9.77 | 1.58 (1.39-1.80) |
| Anaphylaxis | 6 / 8 (2 / 4) | Ondansetron, 3.16; paclitaxel, 0.22 | 1.23 (0.97-1.57) |
| Vomiting | 13 / 56 (11 / 2) | Gabapentin, 5.65; lenvatinib, 0.47 | 1.37 (1.26-1.48) |

Abbreviations: AGEP, acute generalized exanthematous pustulosis; DILI, drug-induced liver injury; DRESS, drug reaction with eosinophilia and systemic symptoms; ICI, immune checkpoint inhibitor; OR, odds ratio; SJS/TEN, Stevens-Johnson syndrome/toxic epidermal necrolysis. Interaction ORs compare the drug-phenotype reporting association in ICI-containing vs non-ICI reports. Time ratios compare documented onset in reports with vs without any-role ICI exposure; values greater than 1 indicate longer onset.

**Figure 3.**
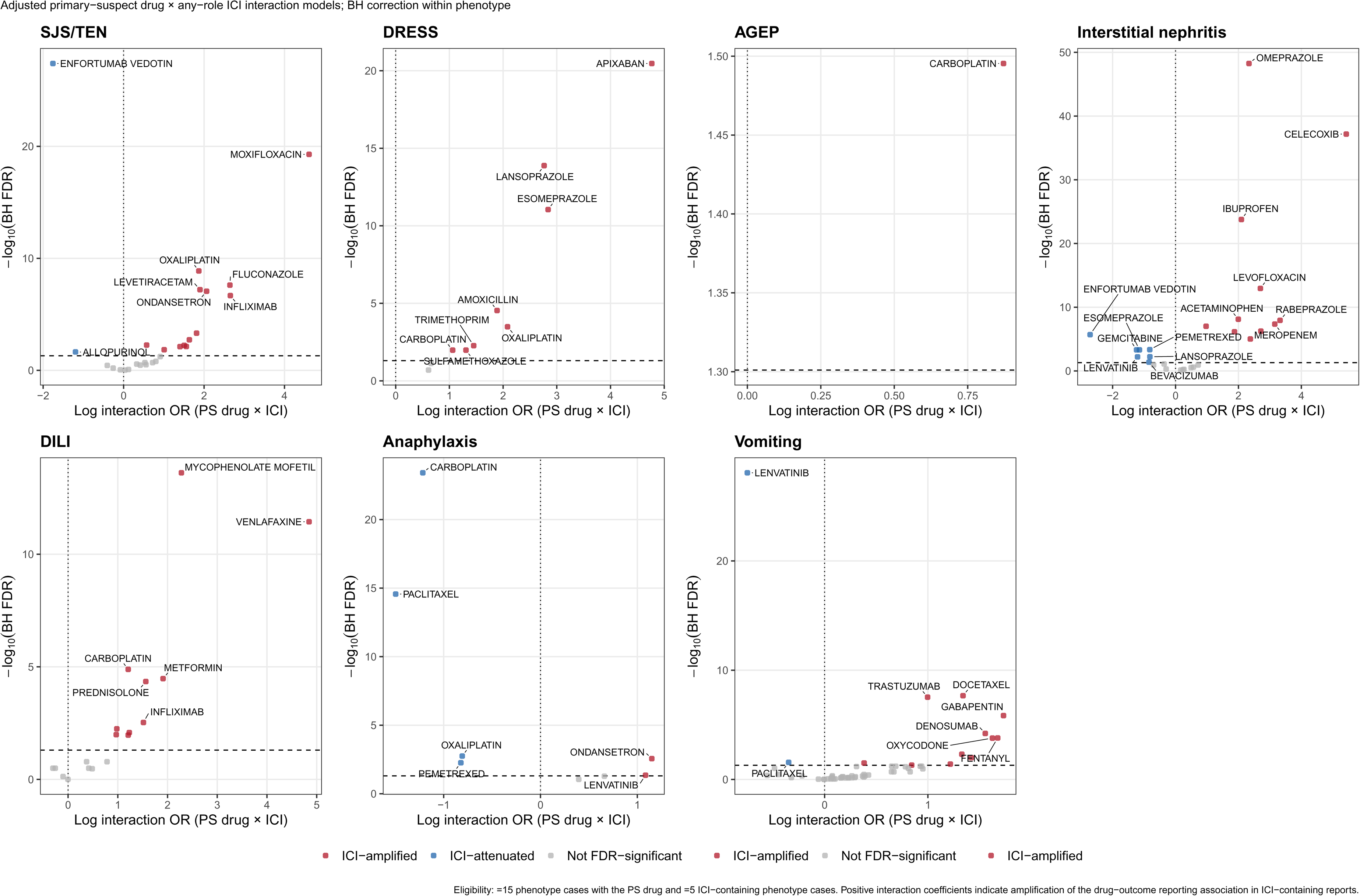
Primary-Suspect Drug × ICI Interactions Are Phenotype-Specific. Adjusted interaction analyses for SJS/TEN, DRESS, AGEP, interstitial nephritis, DILI, anaphylaxis, and vomiting among 13,701,106 FAERS reports. Each point represents an eligible non-ICI primary-suspect drug ingredient with at least 15 phenotype reports overall and at least 5 ICI-containing phenotype reports. Positive interaction coefficients indicate amplification of the drug-phenotype reporting association in ICI-containing reports; negative values indicate attenuation. Models adjusted for age, original age missingness, sex, region, calendar year, number of reported drugs, and cancer status. FDR correction was performed separately within phenotype.

**Figure 4.**
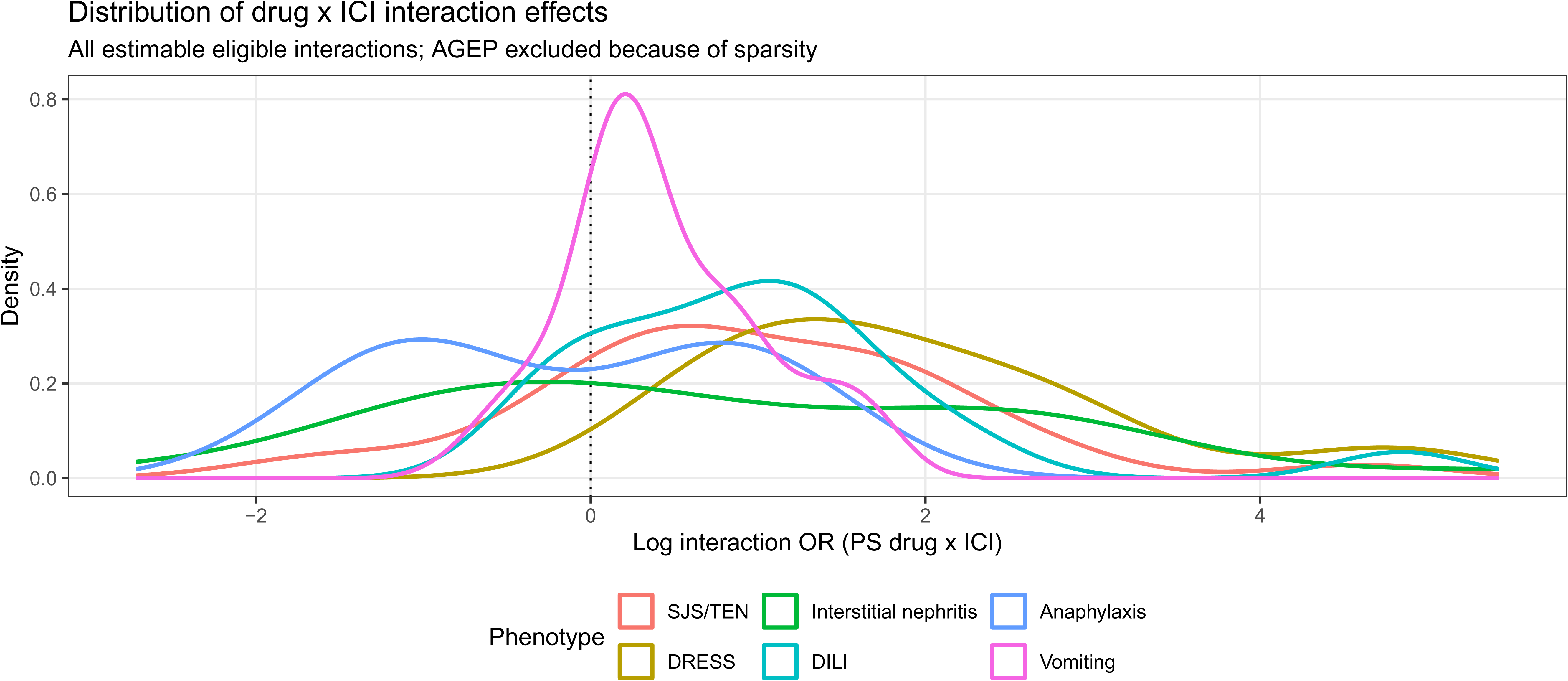
Interaction-Effect Distributions Differ by Phenotype. Density of log interaction ORs across all eligible estimable drug-phenotype pairs, stratified by phenotype. AGEP is omitted because only one drug pair met eligibility criteria. The vertical line at zero denotes no interaction on the log scale.

For AGEP, the single eligible pair, carboplatin, was amplified (interaction OR, 2.39; 95% CI, 1.08-5.30; 13 jointly exposed cases). For DILI, 10 of 18 eligible interactions were significant and all 10 were amplified. The largest estimate involved venlafaxine (127.31; 95% CI, 34.38-471.48; 5 jointly exposed cases); signals with greater joint support included mycophenolate mofetil (9.77; 95% CI, 5.58-17.08; 15 cases) and metformin (6.75; 95% CI, 2.93-15.58; 6 cases). Anaphylaxis had 6 significant interactions among 8 eligible pairs, including 2 amplified and 4 attenuated interactions. Paclitaxel (0.22; 95% CI, 0.16-0.32; 32 jointly exposed cases) and carboplatin (0.30; 95% CI, 0.24-0.37; 93 cases) were attenuated, whereas ondansetron was amplified (3.16; 95% CI, 1.56-6.38; 9 cases). Vomiting had 13 significant interactions among 56 eligible pairs, including 11 amplified and 2 attenuated interactions. Gabapentin (5.65; 95% CI, 2.99-10.69; 11 cases) and fentanyl (5.34; 95% CI, 2.49-11.46; 8 cases) were amplified, whereas lenvatinib was attenuated (0.47; 95% CI, 0.42-0.54; 357 cases).

Across SJS/TEN, DRESS, AGEP, interstitial nephritis, and DILI, 52 of 83 eligible pairs were significant. Interaction-effect distributions were right-shifted for SJS/TEN, DRESS, interstitial nephritis, and DILI, left-shifted for anaphylaxis, and closer to the null for vomiting; AGEP was excluded from the density display because only 1 pair was eligible (**Figure 4**). Of the 71 significant interactions, 61 remained FDR significant in complete-case age analyses and 70 retained the same direction (**eFigure 5**). Median directional stability across 5000 repeated 80% subsamples was 98.5% (IQR, 75.6%-100%); 46 signals retained direction in at least 90% of subsamples and 52 in at least 80% (**eTable 11**). **Figure 5** relates jointly exposed phenotype-case support to coefficient stability, a complementary measure of estimate dispersion. The lowest directional recovery was observed for several signals including vomiting-gemcitabine, SJS/TEN-prednisone, and interstitial nephritis-pemetrexed.

**Figure 5.**
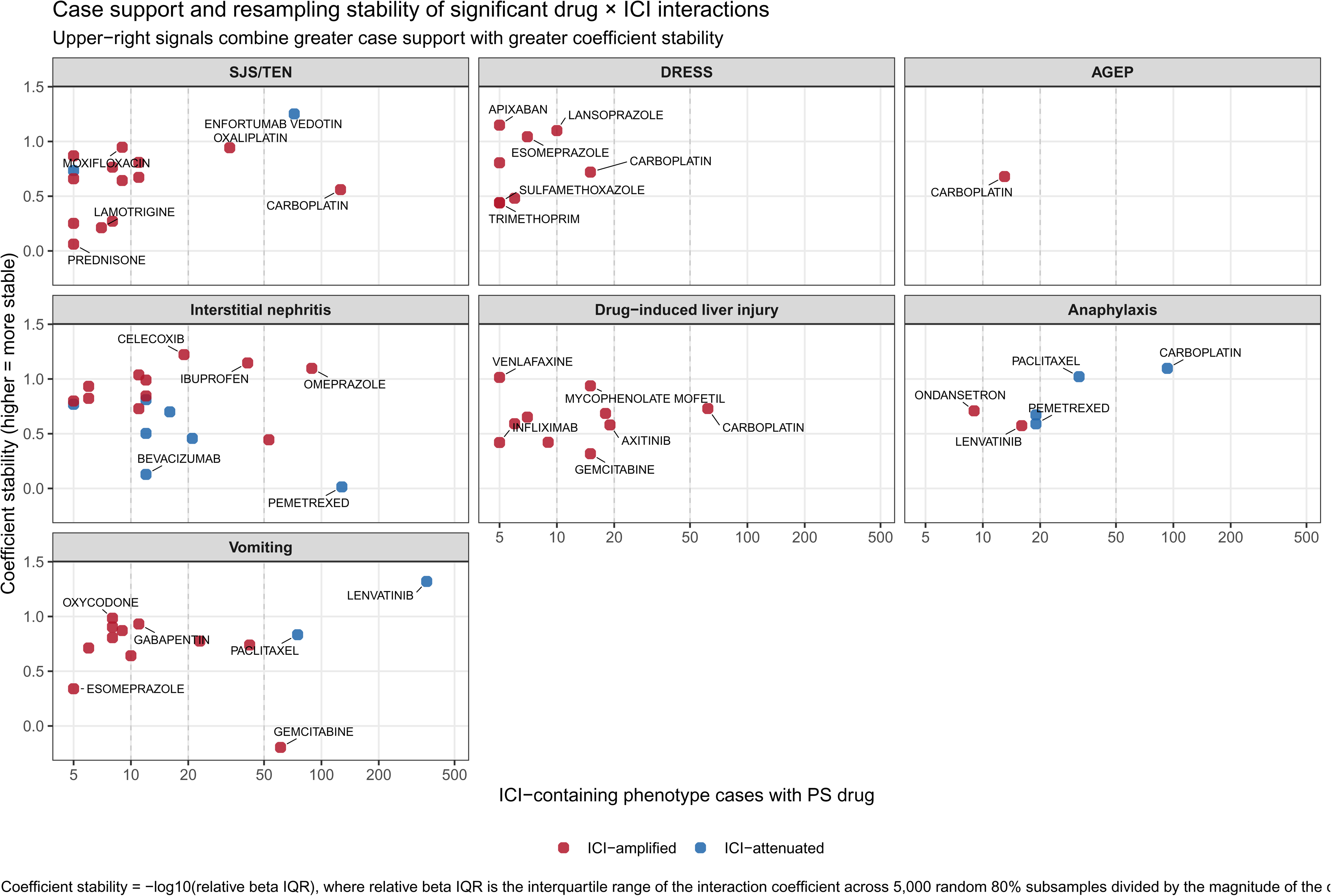
Evidence-Stability Map of Drug × ICI Interactions. Jointly exposed phenotype-case support versus coefficient stability for FDR-significant drug × ICI interactions. Coefficient stability was defined as -log10(relative beta IQR), where relative beta IQR was the interquartile range of the crude interaction coefficient across 5000 random 80% subsamples divided by the absolute value of the observed crude interaction coefficient. Upper-right signals combine greater case support with greater stability.

### Documented onset differs by ICI exposure

A total of 64,593 unique reports contributed to the primary 1- to 365-day temporal cohort. Median documented onset was 20 days with ICI exposure vs 13 days without ICI exposure for SJS/TEN, 49.5 vs 14.5 days for interstitial nephritis, 29 vs 19 days for DILI, and 26 vs 16 days for vomiting. For DRESS, the unadjusted median was 17 days with ICI exposure vs 21 days without ICI exposure; the ICI-exposed distribution had a wider upper tail, while the covariate-adjusted association was in the opposite direction. The temporal comparisons included 692 ICI-exposed and 3439 non-ICI SJS/TEN reports, 133 and 4376 DRESS reports, 528 and 1164 interstitial nephritis reports, 684 and 4227 DILI reports, and 2117 and 42,048 vomiting reports (Table 1; eTable 12). After adjustment, ICI exposure was associated with longer documented onset from the selected primary-suspect treatment start for 5 of 7 phenotypes (**Figure 6**): SJS/TEN (time ratio, 1.37; 95% CI, 1.21-1.54), DRESS (1.43; 95% CI, 1.21-1.69), interstitial nephritis (1.51; 95% CI, 1.19-1.90), DILI (1.58; 95% CI, 1.39-1.80), and vomiting (1.37; 95% CI, 1.26-1.48). AGEP (0.99; 95% CI, 0.67-1.45) and anaphylaxis (1.23; 95% CI, 0.97-1.57) were not significant. Removing the 365-day upper bound retained significant associations for SJS/TEN (1.44; 95% CI, 1.25-1.65) and DILI (1.55; 95% CI, 1.34-1.79). The interstitial nephritis estimate remained similar (1.42; 95% CI, 1.05-1.92) but narrowly missed the FDR threshold (FDR = .051) (**eFigure 6**).

**Figure 6.**
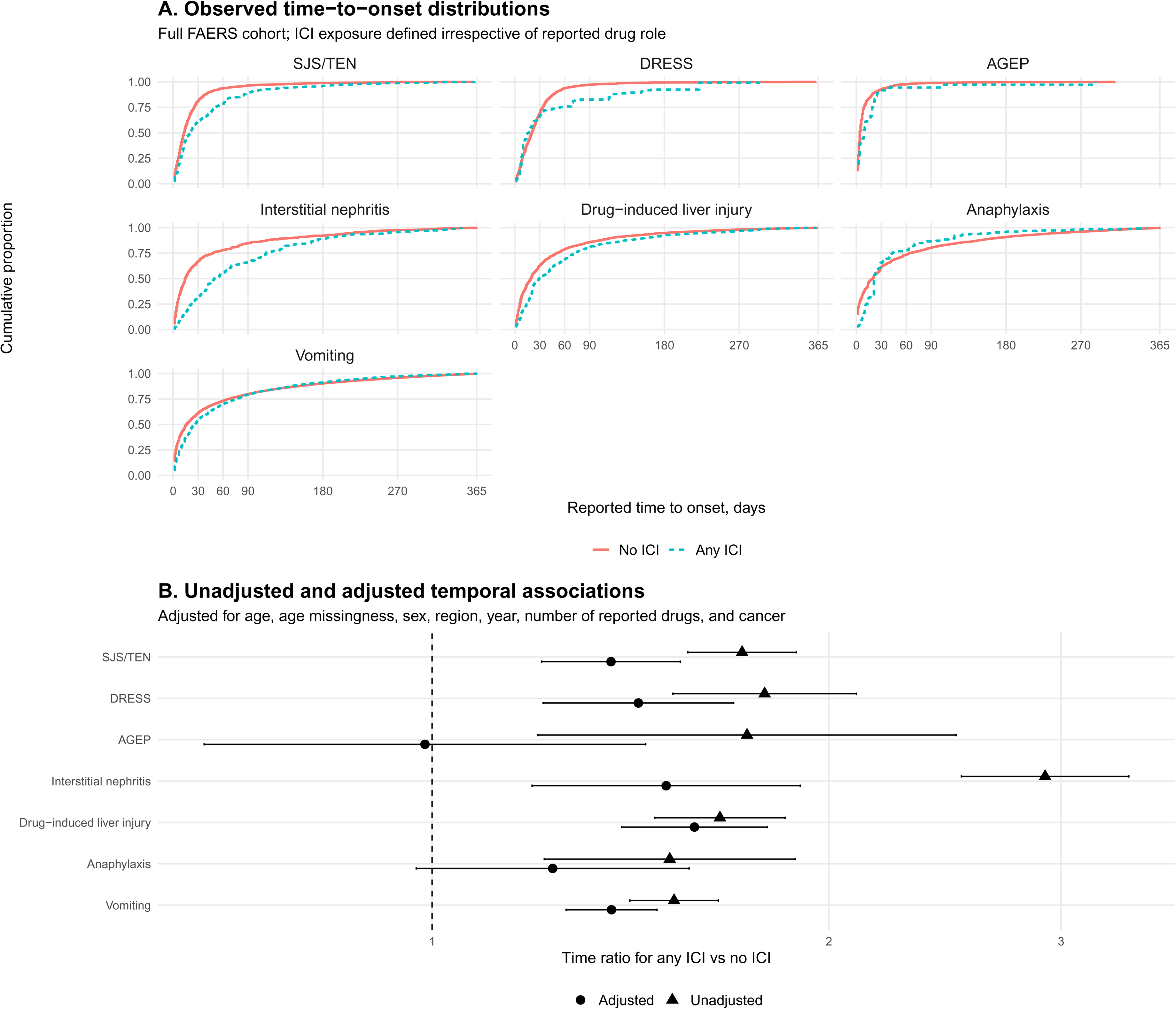
Documented Time to Onset According to ICI Exposure. Temporal analyses among phenotype-positive reports with complete event and primary-suspect treatment-start dates and documented onset of 1 to 365 days. Panel A shows empirical cumulative distributions for SJS/TEN, DRESS, AGEP, interstitial nephritis, DILI, anaphylaxis, and vomiting according to any-role ICI exposure. Panel B shows unadjusted and adjusted accelerated failure-time model estimates. The temporal anchor was the closest positive primary-suspect treatment start preceding the event and does not necessarily represent ICI initiation. Time ratios greater than 1 indicate longer documented time to onset. Adjusted models included age, original age missingness, sex, region, calendar year, number of reported drugs, and cancer status.

### Documented onset also differs by checkpoint pathway

Nineteen checkpoint pathway-phenotype coefficients met estimability criteria; 10 remained significant after global FDR correction (**Figure 7; eTable 13**). PD-1 exposure was associated with longer onset for SJS/TEN (time ratio, 1.38; 95% CI, 1.21-1.57; 605 pathway-positive reports), DRESS (1.52; 95% CI, 1.23-1.87; 108 reports), interstitial nephritis (1.62; 95% CI, 1.27-2.07; 420 reports), DILI (1.26; 95% CI, 1.08-1.49; 422 reports), and vomiting (1.35; 95% CI, 1.23-1.49; 1586 reports). PD-L1 exposure was associated with longer SJS/TEN (1.54; 95% CI, 1.18-1.99; 85 reports), DILI (1.83; 95% CI, 1.52-2.22; 258 reports), and anaphylaxis onset (1.74; 95% CI, 1.18-2.55; 84 reports). CTLA-4 inhibition was associated with shorter interstitial nephritis onset (0.59; 95% CI, 0.41-0.84; 70 reports) and longer DILI onset (1.35; 95% CI, 1.07-1.70; 176 reports). Estimable CTLA-4 associations for SJS/TEN, DRESS, anaphylaxis, and vomiting were not significant, nor were PD-L1 associations for DRESS, interstitial nephritis, and vomiting. AGEP had only 1 estimable pathway coefficient, for PD-1, which was not significant. LAG-3, TIGIT, and TIM-3 exposure was too sparse for phenotype-specific estimation.

**Figure 7.**
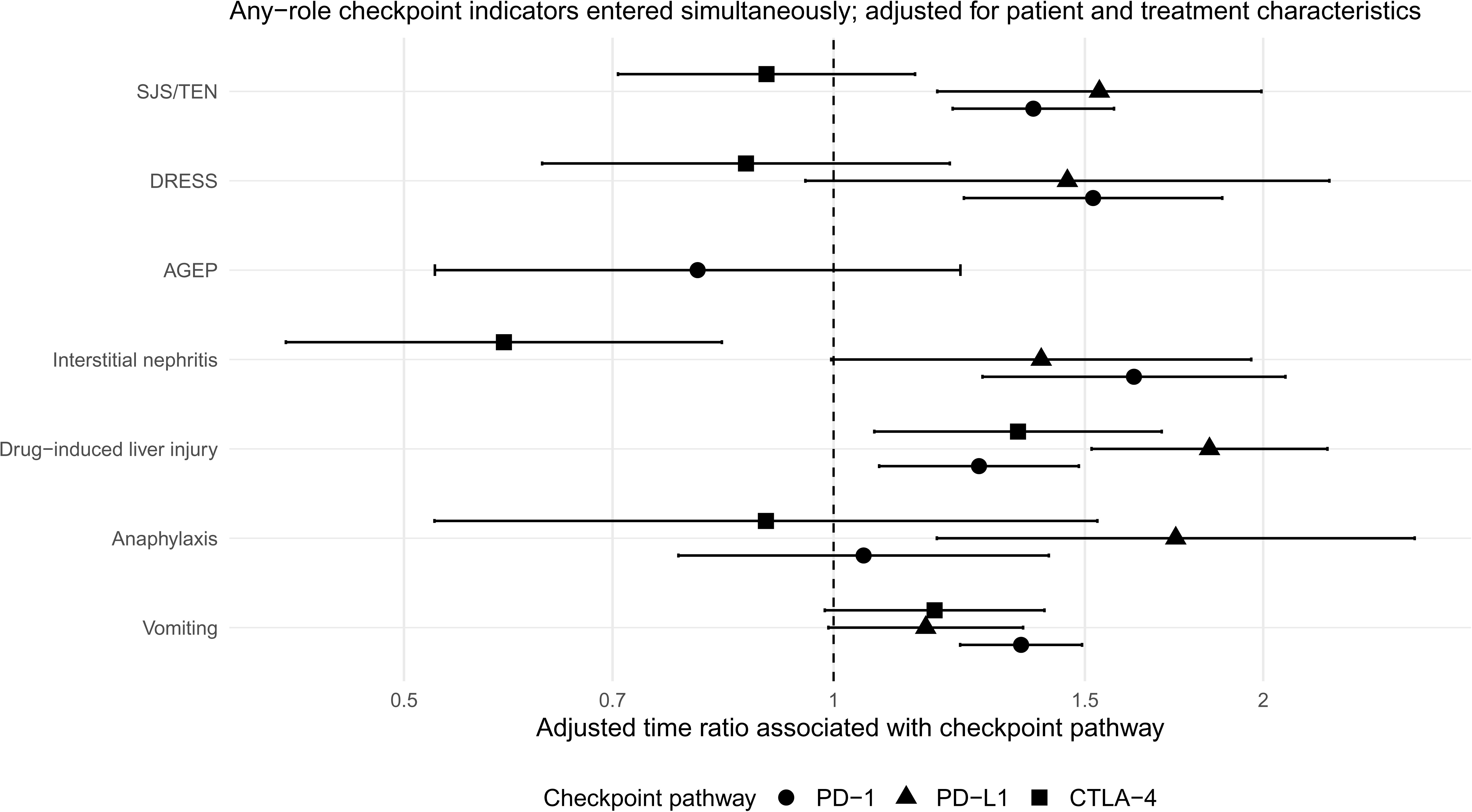
Checkpoint Pathway-Specific Temporal Associations. Adjusted accelerated failure-time models evaluating PD-1, PD-L1, CTLA-4, LAG-3, TIGIT, and TIM-3 pathway exposure using the Figure 6 temporal substrate. Supported checkpoint indicators were entered simultaneously within each phenotype-specific model; combination regimens could therefore contribute to multiple pathway coefficients. Nineteen coefficients were estimable; LAG-3, TIGIT, and TIM-3 did not meet support requirements for phenotype-specific estimation. Time ratios greater than 1 indicate longer documented time to onset associated with a pathway-positive vs pathway-negative report, conditional on the other supported pathway indicators and covariates. FDR correction was applied across all estimable checkpoint-phenotype coefficients.

## Discussion

The central finding is a change in relationships, not simply an increase in toxicity. Across 13.7 million reports, checkpoint blockade was associated with an altered treatment state in which the reported effects of other drugs depended on the phenotype and checkpoint context. In a mechanistic two-hit model, the ICI need not be the final tissue-damaging exposure: the first hit may change immune permissiveness, while a second medication helps determine whether and how toxicity is expressed. That state left several independent signatures – drug-phenotype associations moved in both directions, the adverse-event landscape was redistributed, toxicities formed cross-organ structure, and documented onset varied by checkpoint pathway. None establishes mechanism alone; together, they define a pattern that is difficult to reduce to a single generic reporting tendency.

The global landscape and network analyses act as stress tests of that interpretation rather than as the primary mechanistic claim. Nearly equal numbers of eligible Preferred Terms had higher and lower reporting odds with ICI exposure, arguing against indiscriminate inflation of adverse-event reporting.

Within ICI-containing reports, the unsupervised network recovered a neuromuscular-cardiac community containing the recognized myocarditis-myositis-myasthenia overlap syndrome^19–20^ without specifying that syndrome in advance. That recovery provides useful face validity, consistent with prior work showing that FAERS co-reporting networks can recover clinically meaningful structure.^24^ The less familiar gastrointestinal-systemic-neurologic, cutaneous-systemic-neurologic and endocrine-vascular-renal communities are better treated as hypotheses about co-occurring clinical states than as new syndromes. Their edges may reflect shared inflammation, diagnostic cascades, treatment complications, or preferential reporting of complex cases. The network does not establish shared pathogenesis – its narrower contribution is to show that the ICI-associated treatment state has structure beyond the individual drug-phenotype pairs that motivated the analysis.

Mechanistically, the shortest path from checkpoint blockade to altered drug toxicity is a lowered threshold for drug-specific T-cell priming. PD-1/PD-L1 blockade can make naive T cells responsive to drug-associated antigens at exposures that would otherwise remain below the priming threshold,^5–6^ while hypersensitivity to previously tolerated contrast media and sequential targeted therapies provides a human analogue.^7–10^ Other checkpoint effects could widen that permissive state through expansion or reinvigoration of T-cell clones, recognition of shared tumor and tissue antigens, early B-cell perturbation, tissue-specific inflammatory programs, and microbiome-associated immune modulation.^12–15,17–18^ These mechanisms are not mutually exclusive. What they share is a falsifiable prediction: checkpoint blockade should amplify some drug-phenotype relationships, leave others comparatively unchanged, and potentially attenuate others when the underlying toxicity is driven by different biology. That is much closer to the pattern observed here than a model in which every co-medication simply inherits the background toxicity of the ICI.

A two-hit model worth taking seriously must make differential predictions; it cannot merely predict that two exposures are worse than one. The interaction screen did exactly that. SJS/TEN, DRESS, interstitial nephritis and DILI shifted predominantly toward amplification, whereas anaphylaxis more often showed attenuation and vomiting remained closer to the null. SJS/TEN and DRESS are drug-specific T-cell-mediated syndromes, while anaphylaxis is rapid and largely mast-cell driven and vomiting is common, nonspecific and tightly coupled to the underlying cancer regimen. Interstitial nephritis and DILI occupy a more heterogeneous middle ground that can include checkpoint autoimmunity, classical drug hypersensitivity, direct toxicity or mixtures of these mechanisms. The left-shifted anaphylaxis distribution and comparatively null vomiting distribution are therefore important negative contrasts: they argue against a trivial process in which any drug recorded beside an ICI acquires an amplified interaction signal.

The individual signals also show why an interaction term cannot be separated from the biology of the drug. Moxifloxacin showed marked SJS/TEN amplification. Fluoroquinolones are established, if uncommon, SJS/TEN culprits,^25^ and a recent report described moxifloxacin-associated TEN after PD-1 blockade with causality assessments implicating moxifloxacin as the principal trigger.^26^ Enfortumab vedotin produced a qualitatively different signal. It has substantial intrinsic cutaneous toxicity and can cause severe and fatal SJS/TEN,^27^ but that injury may not depend on the same checkpoint-enabled hypersensitivity mechanism. ICI-associated epidermal necrolysis itself can show a strongly inflammatory immune program, including macrophage-derived CXCL10,^28^ whereas enfortumab vedotin may produce on-target cutaneous injury through nectin-4 expression in the skin.^27^ Combination studies reported more frequent cutaneous adverse events with enfortumab vedotin plus pembrolizumab, but these were broad skin-toxicity categories rather than specifically desquamative reactions or SCAR.^29^ Earlier enfortumab studies also enrolled patients after PD-1/PD-L1 therapy, so FAERS reports without a concurrently listed ICI may still include remote checkpoint exposure.^30^ The submultiplicative interaction can therefore coexist with substantial severe cutaneous risk: it may reflect strong independent enfortumab toxicity without additional checkpoint-mediated amplification, compounded by incomplete exposure history, rather than protection.

DRESS produced perhaps the cleanest phenotype-level amplification pattern: eight of ten eligible interactions were significant, and every one pointed in the same direction. That coherence is striking for a syndrome fundamentally driven by drug-specific T-cell responses. Several amplified drugs already have independent evidence as DRESS culprits. Proton pump inhibitor-associated DRESS has been reported across esomeprazole, pantoprazole, omeprazole and lansoprazole, including an omeprazole case supported by drug-specific ELISpot testing;^31^ apixaban-associated DRESS has also been reported.^32^ The more interesting interpretation is therefore not that checkpoint blockade creates an entirely new set of culprit drugs, but that it may amplify pre-existing hypersensitivity liability. The apixaban estimate remains imprecise because only five jointly exposed cases supported it, but its direction is concordant with the broader DRESS pattern and with experimental evidence that PD-1/PD-L1 blockade lowers the threshold for drug-specific T-cell priming.^5–6^

The omeprazole-interstitial nephritis interaction is one of the strongest signals with external clinical coherence. Proton pump inhibitor exposure repeatedly appears among patients with ICI-associated acute kidney injury, and acute tubulointerstitial nephritis is the dominant biopsy pattern in reported series.^33–36^ A biologically attractive explanation is that checkpoint blockade releases drug-specific T cells previously held in check during exposure to a medication capable of causing interstitial nephritis on its own. FAERS cannot distinguish that mechanism from alternatives: proton pump inhibitors are common in patients with cancer, may mark comorbidity or healthcare exposure, and may be assigned different suspect roles when an ICI is present. The signal therefore argues for a denominator-based longitudinal cohort with medication timing and adjudicated kidney injury – not immediate avoidance of a widely used drug class.

The altered treatment state also appeared to change the clock. ICI-containing reports had longer documented onset from the selected primary-suspect drug start for five of seven phenotypes in the prespecified 1-365-day analysis. This is deliberately different from the usual question of latency after ICI initiation: it asks whether a drug-associated event unfolds on a different timescale when checkpoint therapy is also present. Longer intervals could reflect immune evolution, sequential exposure, or altered attribution, and the weakening of several estimates after removing the 365-day ceiling shows how strongly the reporting tail can matter. Delayed irAEs after treatment discontinuation are recognized, and organ-specific onset varies substantially in trial and observational data.^37–39^ Pathway models added another layer: interstitial nephritis had opposing PD-1 and CTLA-4 associations, whereas DILI onset was longer with PD-1, PD-L1 and CTLA-4 exposure. Distinct checkpoint biology is plausible, but these coefficients also encode the tumors, combinations and treatment lines in which each pathway is used.^40–41^

Clinically, the two-hit framework argues for relational rather than single-drug culprit assessment. When a patient receiving an ICI develops hepatitis, nephritis or a severe eruption, the checkpoint inhibitor is the obvious suspect – but it may not be the only relevant exposure. Exclusive attribution can obscure a recently added antimicrobial, acid suppressant or targeted therapy; the converse is equally important, because a large FAERS interaction OR is not sufficient to label a co-medication unsafe. Medication start dates, prior tolerance, dechallenge, rechallenge, phenotype-specific latency and alternative causes should be reviewed alongside established irAE algorithms.^3–4,42^ This matters most when attribution determines permanent ICI discontinuation, high-dose immunosuppression or avoidance of a medication important for cancer care.

The atlas is therefore a map of mechanistic priorities, not a prescribing table. High-support signals with external coherence, such as proton pump inhibitors and interstitial nephritis, can move first into longitudinal oncology cohorts with treatment denominators, time-varying exposures, tumor type and stage, regimen, renal or hepatic function and adjudicated outcomes. Large estimates supported by few joint cases require replication before mechanistic investment. Validated pairs can then be interrogated with HLA typing, drug-specific T-cell assays, tissue profiling and perturbation models to separate checkpoint-enabled loss of tolerance from direct injury or confounding by treatment ecology. That sequence – from interaction signal, to longitudinal validation, to mechanism – is the most direct test of whether the two-hit model is biologically real.

## Limitations

FAERS is a spontaneous-reporting system without treatment denominators; reporting ORs cannot estimate incidence, absolute risk or causal effects.^22–23,43–44^ Reports are influenced by publicity, indication, disease severity, treatment era, surveillance and polypharmacy. Drug-role assignment is supplied by the reporter and may be particularly unreliable for medications started to treat early symptoms of an evolving toxicity, creating protopathic or reverse-causation signals. The primary-suspect restriction improves phenotype attribution but can also exclude true contributors assigned another role. Conversely, role-agnostic ICI exposure maximizes capture of checkpoint therapy but cannot determine whether the ICI preceded the drug or event in every report. The interaction term is therefore a measure of departure from multiplicative reporting odds, not biological synergy. Cancer ascertainment from indications and treatment proxies is imperfect, and residual differences in tumor type, stage, treatment line, regimen, performance status and supportive care may remain despite covariate adjustment. These factors are particularly relevant to checkpoint-pathway comparisons because pathway use is not exchangeable across cancers or eras.

Although the source data were deduplicated, duplicate reporting remains a recognized challenge in FAERS.^45^ Some interaction estimates were based on small numbers of jointly exposed cases and consequently had wide CIs; repeated subsampling assesses internal directional stability but does not correct bias or provide external replication. Age was frequently missing and required imputation, although complete-case estimates were highly directionally concordant. The temporal analyses selected the closest preceding exact-date primary-suspect start and excluded partial dates, so they describe documented reporting intervals in a selected subset rather than prospective latency from ICI initiation. The 365-day primary window influenced some estimates, as shown by the unrestricted sensitivity analysis. Co-reporting networks are also descriptive: edges can arise from shared pathogenesis, diagnostic cascades, treatment of one event leading to another, or preferential reporting of complex cases. Finally, multiple-testing control limits false discoveries within the specified analyses but does not remove selection, measurement or reporting bias. These limitations make the findings appropriate for signal prioritization and hypothesis testing, not direct prescribing rules.

## Conclusions

Immune checkpoint blockade was associated with more than an additional layer of toxicity: it changed the reported relationships between other drugs and specific adverse phenotypes, while toxicity organization and documented onset varied with checkpoint exposure. These convergent findings support a two-hit treatment-state model in which ICIs may reset immune permissiveness and thereby change the toxicity liability of subsequent or concomitant medications. Longitudinal and mechanistic validation is needed before individual signals can guide prescribing, but the framework provides a testable way to ask how cancer immunotherapy reshapes drug tolerance.

## Data Availability

All data produced in the present study are available upon reasonable request to the authors

## Data availability statement

Source FAERS data are publicly available from the US Food and Drug Administration. Curated analysis dictionaries, derived result tables and reproducible analysis code will be made available in a public repository at publication.

## Contributors

EM and EP conceived the study. EM, AA, CAS, MM-P and EP designed the analyses. EM curated the data and performed the analyses with statistical input from AA. DP, MSK, CAS, MM-P and EP contributed to clinical interpretation. EM drafted the manuscript, and all authors critically revised and approved the final version. EM had full access to the data, accepts responsibility for the integrity and accuracy of the analysis and is the guarantor.

## Ethics approval

Not applicable. FAERS contains publicly available, deidentified reports; institutional review board approval was not required.

## Patient consent for publication

Not applicable.

## Patient and public involvement

Patients and/or the public were not involved in the design, conduct, reporting or dissemination plans of this research.

## Artificial intelligence use

OpenAI GPT-5.6 Sol was used to assist with candidate drug-name normalization (as detailed in the Online Supplement), R code development and debugging, idea generation, and manuscript drafting and editing. All drug mappings, code, analyses, statistical outputs, citations, and manuscript text were manually reviewed and verified by the authors, who take responsibility for the integrity and accuracy of the work.

## Supplement 1. Supplementary Methods, Tables, and Figures

eMethods. Detailed FAERS import and deduplication, canonical drug mapping, GPT-5.6 Sol-assisted candidate assignment and manual adjudication, cohort construction, exposure and phenotype definitions, missing-age imputation, network construction, interaction modeling, temporal modeling, software packages, and sensitivity analyses.

eTable 1. Cohort Flow Counts for Main and Temporal Analyses.

eTable 2. Checkpoint Inhibitor Agent Reports by Year, 2016-2025.

eTable 3. Checkpoint Target Reports by Year, 2016-2025.

eTable 4. Checkpoint Inhibitor Dictionary and Target Mapping.

eTable 5. Complete Cancer-Restricted Preferred-Term Association Results Underlying Figure 1.

eTable 6. Cross-Organ HLT Community Membership Underlying Figure 2.

eTable 7. Cross-Organ Community Audit Metrics Underlying Figure 2.

eTable 8. Primary HLT Edges Retained in Figure 2.

eTable 9. Figure 2 Edge-Threshold Sensitivity Analysis.

eTable 10. Complete Primary-Suspect Drug × ICI Interaction Results Underlying Figure 3.

eTable 11. Interaction Robustness, Directional Stability, and Coefficient Stability Underlying Figures 3 and 5.

eTable 12. Figure 6 Temporal Summary, AFT Results, and Unrestricted-Time Sensitivity.

eTable 13. Complete Figure 7 Checkpoint-Pathway Temporality Results.

**eFigure 1.**
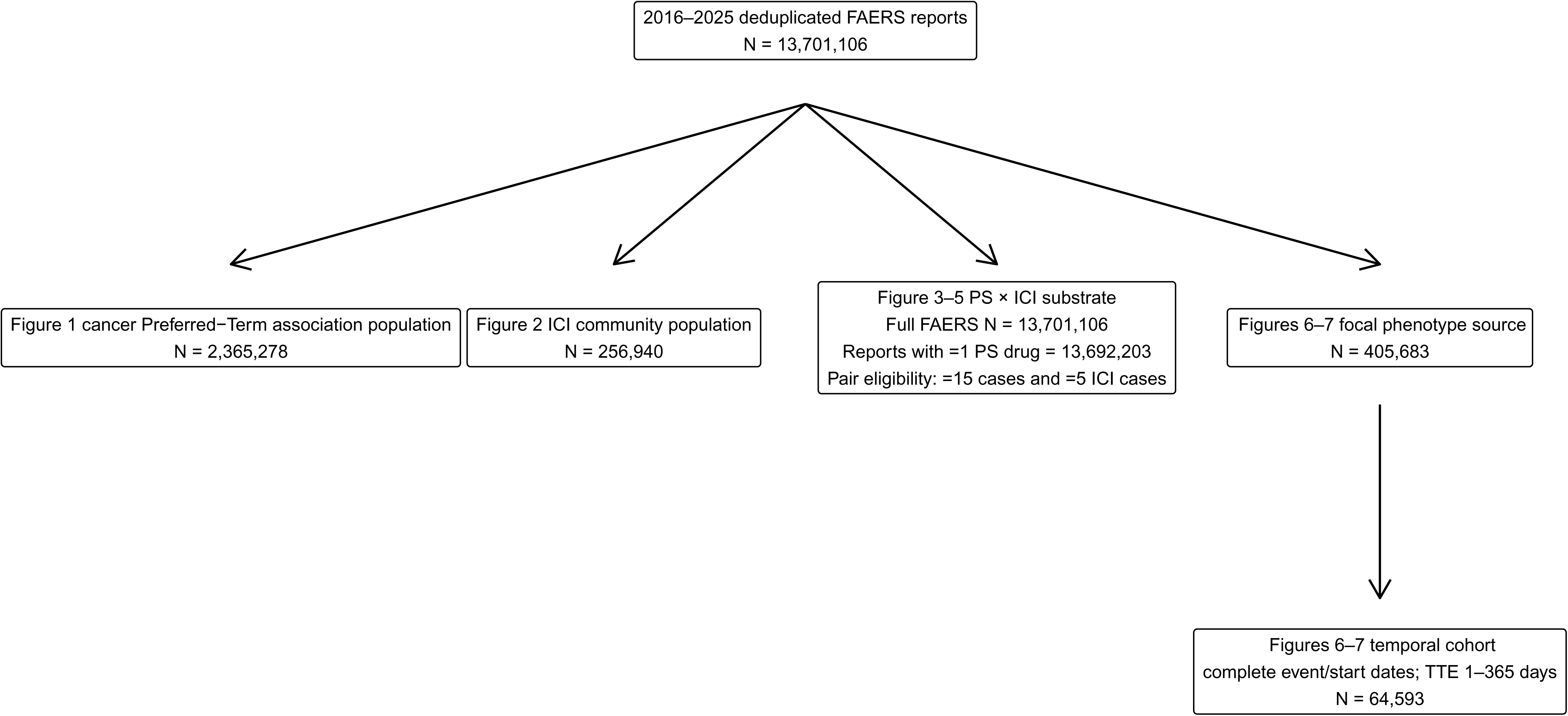
Cohort and Analysis Flow Diagram.

**eFigure 2.**
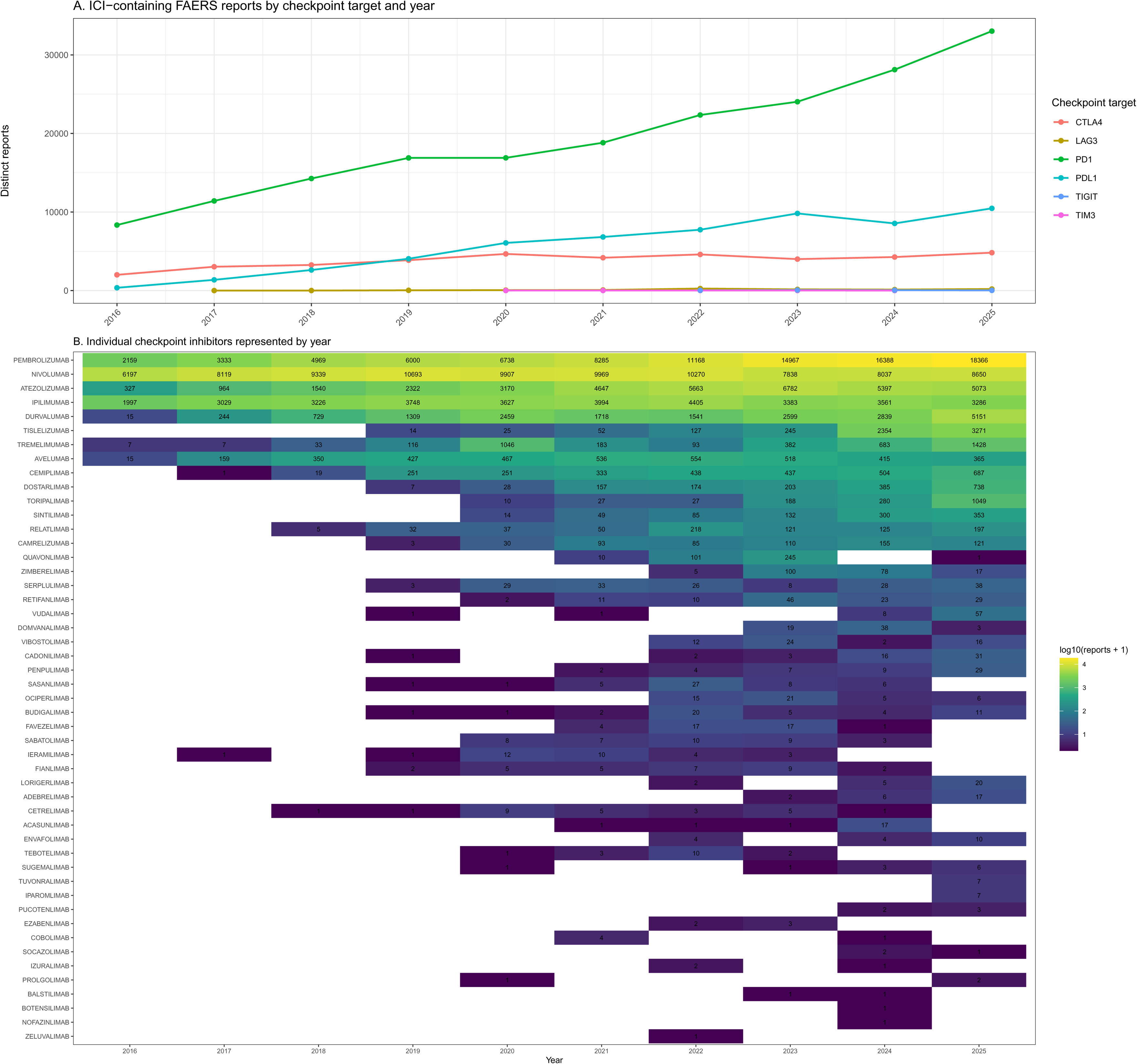
Checkpoint Inhibitor Agents and Reports by Year.

**eFigure 3.**
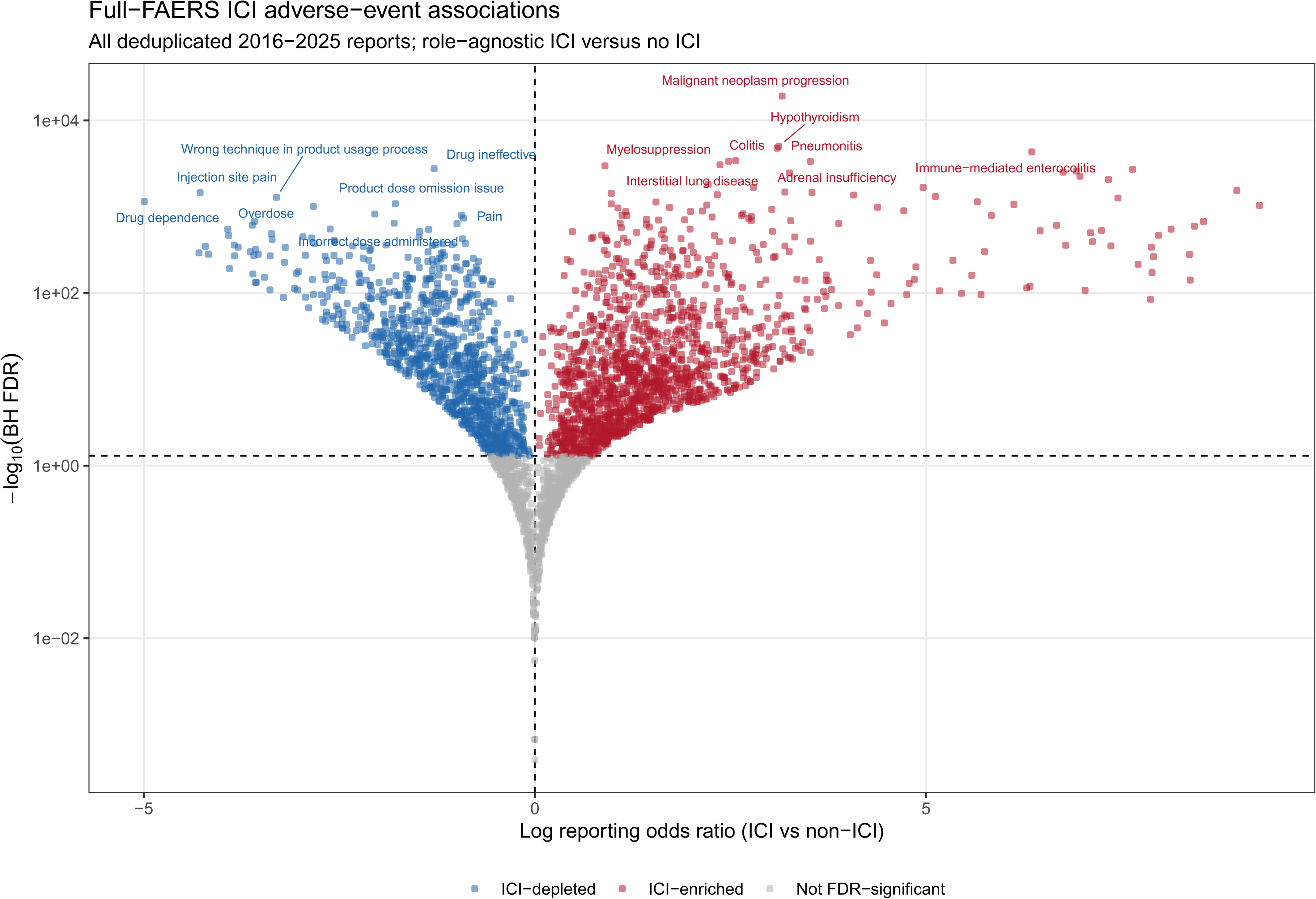
Full-FAERS Preferred-Term Association Sensitivity Analysis.

**eFigure 4.**
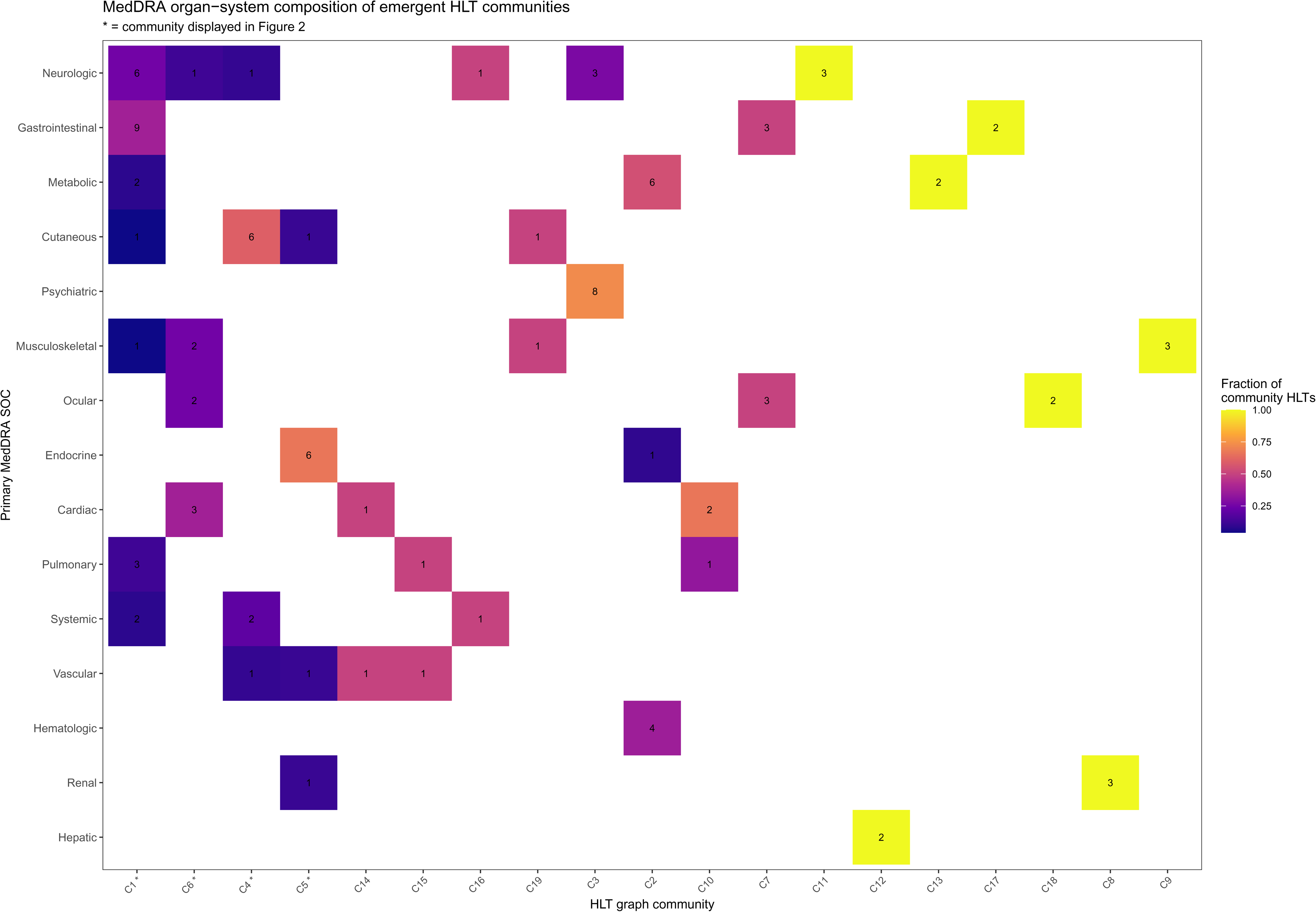
Community-by-System-Organ-Class Heatmap.

**eFigure 5.**
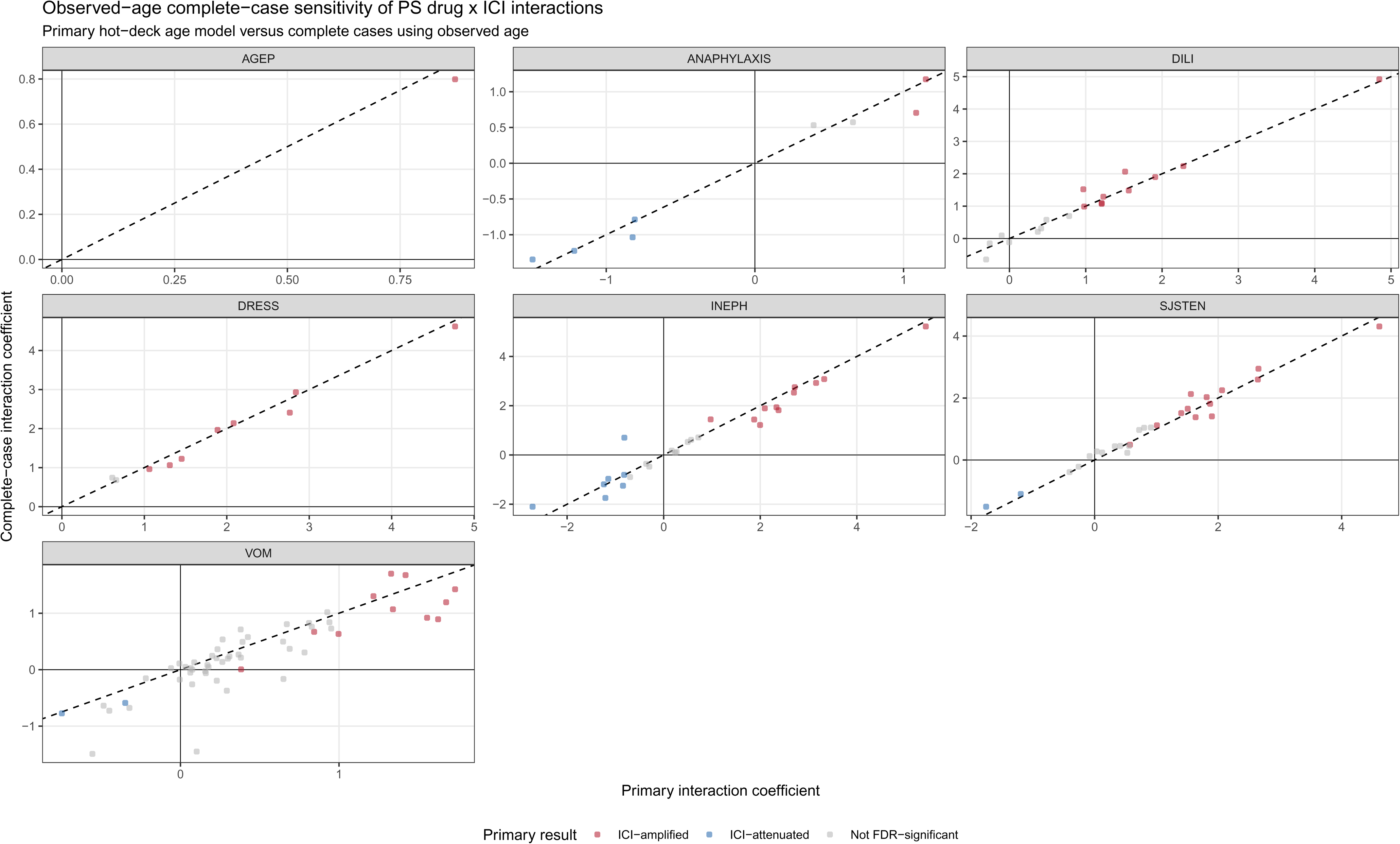
Complete-Case Age Sensitivity Analysis for Drug × ICI Interactions.

**eFigure 6.**
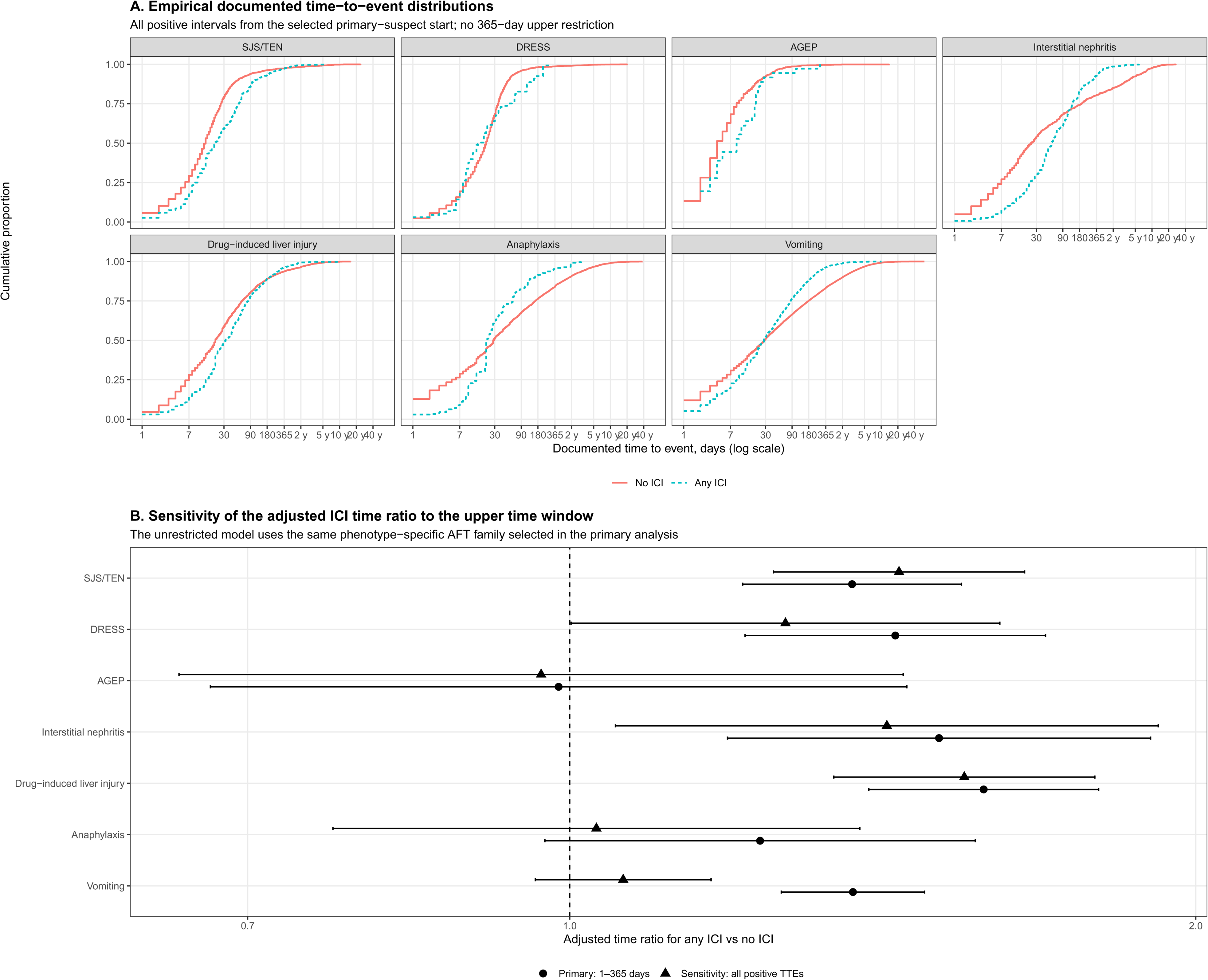
Documented Time-to-Event Distributions and Sensitivity to the 365-Day Analysis Window.

## References

1. Topalian SL, Hodi FS, Brahmer JR, et al. Five-Year Survival and Correlates Among Patients With Advanced Melanoma, Renal Cell Carcinoma, or Non-Small Cell Lung Cancer Treated With Nivolumab. JAMA Oncol. 2019;5(10):1411–1420. doi:10.1001/jamaoncol.2019.2187

2. Wei SC, Duffy CR, Allison JP. Fundamental Mechanisms of Immune Checkpoint Blockade Therapy. Cancer Discovery. 2018;8(9):1069–1086. doi:10.1158/2159-8290.CD-18-0367

3. Schneider BJ, Naidoo J, Santomasso BD, et al. Management of Immune-Related Adverse Events in Patients Treated With Immune Checkpoint Inhibitor Therapy: ASCO Guideline Update. J Clin Oncol. 2021;39(36):4073–4126. doi:10.1200/JCO.21.01440

4. Brahmer JR, Abu-Sbeih H, Ascierto PA, et al. Society for Immunotherapy of Cancer (SITC) clinical practice guideline on immune checkpoint inhibitor-related adverse events. J Immunother Cancer. 2021;9(6):e002435. doi:10.1136/jitc-2021-002435

5. Grice S, Saide K, Farrell L, et al. Immune checkpoint blockade lowers the threshold of naïve T-cell priming to drug-associated antigens in a dose-dependent fashion. Toxicol Sci. 2024;202(1):13–18. doi:10.1093/toxsci/kfae118

6. Hammond S, Olsson-Brown A, Grice S, et al. Checkpoint Inhibition Reduces the Threshold for Drug-Specific T-Cell Priming and Increases the Incidence of Sulfasalazine Hypersensitivity. Toxicological Sciences. 2022;186(1):58–69. doi:10.1093/toxsci/kfab144

7. Hammond S, Olsson-Brown A, Gardner J, et al. T cell mediated hypersensitivity to previously tolerated iodinated contrast media precipitated by introduction of atezolizumab. Journal for ImmunoTherapy of Cancer. 2021;9(5):e002521. doi:10.1136/jitc-2021-002521

8. Rana J, Maloney NJ, Rieger KE, et al. Drug-induced hypersensitivity syndrome like reaction with angioedema and hypotension associated with BRAF inhibitor use and antecedent immune checkpoint therapy. JAAD Case Reports. 2021;13:147–151. doi:10.1016/j.jdcr.2021.04.033

9. Maloney NJ, Rana J, Yang JJ, Zaba LC, Kwong BY. Clinical features of drug-induced hypersensitivity syndrome to BRAF inhibitors with and without previous immune checkpoint inhibition: a review. Supportive Care in Cancer. 2022;30(3):2839–2851. doi:10.1007/s00520-021-06543-9

10. Naqash AR, File DM, Ziemer CM, et al. Cutaneous adverse reactions in B-RAF positive metastatic melanoma following sequential treatment with B-RAF/MEK inhibitors and immune checkpoint blockade or vice versa. A single-institutional case-series. Journal for ImmunoTherapy of Cancer. 2019;7(1):4. doi:10.1186/s40425-018-0475-y

11. Mukherjee EM, Park D, Asiaee A, et al. Immune Checkpoint Inhibitors as Independent and Synergistic Drivers of SJS/TEN. JAMA Oncol. 2025;11(12):1542–1545. doi:10.1001/jamaoncol.2025.4349

12. Das R, Bar N, Ferreira M, et al. Early B cell changes predict autoimmunity following combination immune checkpoint blockade. J Clin Invest. 2018;128(2):715–720. doi:10.1172/JCI96798

13. Lozano AX, Chaudhuri AA, Nene A, et al. T cell characteristics associated with toxicity to immune checkpoint blockade in patients with melanoma. Nat Med. 2022;28(2):353–362. doi:10.1038/s41591-021-01623-z

14. Berner F, Bomze D, Diem S, et al. Association of Checkpoint Inhibitor-Induced Toxic Effects With Shared Cancer and Tissue Antigens in Non-Small Cell Lung Cancer. JAMA Oncol. 2019;5(7):1043–1047. doi:10.1001/jamaoncol.2019.0402

15. Luoma AM, Suo S, Williams HL, et al. Molecular Pathways of Colon Inflammation Induced by Cancer Immunotherapy. Cell. 2020;182(3):655–671.e22. doi:10.1016/j.cell.2020.06.001

16. Jing Y, Chen X, Li K, et al. Association of antibiotic treatment with immune-related adverse events in patients with cancer receiving immunotherapy. Journal for ImmunoTherapy of Cancer. 2022;10(1):e003779. doi:10.1136/jitc-2021-003779

17. McCulloch JA, Davar D, Rodrigues RR, et al. Intestinal microbiota signatures of clinical response and immune-related adverse events in melanoma patients treated with anti-PD-1. Nat Med. 2022;28(3):545–556. doi:10.1038/s41591-022-01698-2

18. Andrews MC, Duong CPM, Gopalakrishnan V, et al. Gut microbiota signatures are associated with toxicity to combined CTLA-4 and PD-1 blockade. Nat Med. 2021;27(8):1432–1441. doi:10.1038/s41591-021-01406-6

19. Wan G, Chen W, Khattab S, et al. Multi-organ immune-related adverse events from immune checkpoint inhibitors and their downstream implications: a retrospective multicohort study. Lancet Oncol. 2024;25(8):1053–1069. doi:10.1016/S1470-2045(24)00278-X

20. Pathak R, Katel A, Massarelli E, Villaflor VM, Sun V, Salgia R. Immune Checkpoint Inhibitor-Induced Myocarditis with Myositis/Myasthenia Gravis Overlap Syndrome: A Systematic Review of Cases. Oncologist. 2021;26(12):1052–1061. doi:10.1002/onco.13931

21. Wang Z, Peng Y, Zhou JG, et al. faers: A High-Fidelity Framework and R/Bioconductor Package for Precision Adverse Event Surveillance. medRxiv. Published online 2026. doi:10.64898/2026.03.26.26349444

22. Bate A, Evans SJW. Quantitative signal detection using spontaneous ADR reporting. Pharmacoepidemiol Drug Saf. 2009;18(6):427–436. doi:10.1002/pds.1742

23. Rothman KJ, Lanes S, Sacks ST. The reporting odds ratio and its advantages over the proportional reporting ratio. Pharmacoepidemiol Drug Saf. 2004;13(8):519–523. doi:10.1002/pds.1001

24. Davis R, Dang O, De S, Ball R. Characterizing the FDA Adverse Event Reporting System (FAERS) as a Network to Improve Pattern Discovery. Drug Safety. 2026;49(2):239–251. doi:10.1007/s40264-025-01609-7

25. Lee EY, Knox C, Phillips EJ. Worldwide Prevalence of Antibiotic-Associated Stevens-Johnson Syndrome and Toxic Epidermal Necrolysis: A Systematic Review and Meta-analysis. JAMA Dermatol. 2023;159(4):384–392. doi:10.1001/jamadermatol.2022.6378

26. Li Q, Wang L, Xu H, et al. Concurrent moxifloxacin-induced liver injury and toxic epidermal necrolysis after immune checkpoint inhibition: a case report and literature review. Front Immunol. 2026;17:1753434. doi:10.3389/fimmu.2026.1753434

27. Viscuse PV, Marques-Piubelli ML, Heberton MM, et al. Case Report: Enfortumab Vedotin for Metastatic Urothelial Carcinoma: A Case Series on the Clinical and Histopathologic Spectrum of Adverse Cutaneous Reactions From Fatal Stevens-Johnson Syndrome/Toxic Epidermal Necrolysis to Dermal Hypersensitivity Reaction. Front Oncol. 2021;11:621591. doi:10.3389/fonc.2021.621591

28. Chen CB, Hung SI, Chang JWC, et al. Immune checkpoint inhibitor-induced severe epidermal necrolysis mediated by macrophage-derived CXCL10 and abated by TNF blockade. Nat Commun. 2024;15(1):10733. doi:10.1038/s41467-024-54180-7

29. O’Donnell PH, Milowsky MI, Petrylak DP, et al. Enfortumab Vedotin With or Without Pembrolizumab in Cisplatin-Ineligible Patients With Previously Untreated Locally Advanced or Metastatic Urothelial Cancer. J Clin Oncol. 2023;41(25):4107–4117. doi:10.1200/JCO.22.02887

30. Rosenberg JE, O’Donnell PH, Balar AV, et al. Pivotal Trial of Enfortumab Vedotin in Urothelial Carcinoma After Platinum and Anti-Programmed Death 1/Programmed Death Ligand 1 Therapy. J Clin Oncol. 2019;37(29):2592–2600. doi:10.1200/JCO.19.01140

31. Pinyopornpanish K, Pinyopornpanish K, Pinyopornpanish K, et al. Omeprazole Induced Rapid Drug Reaction with Eosinophilia, Systemic Symptoms, and Cross-Reactivity in Delayed-Type Hypersensitivity Associated with Proton-Pump Inhibitors: A Case Report and Literature Review. Case Rep Immunol. 2024;2024:1317971. doi:10.1155/2024/1317971

32. Menteşoğlu D, Doğan Günaydın S, Ersoy-Evans S. Drug reaction with eosinophilia and systemic symptoms syndrome induced by apixaban. Dermatol Ther. 2020;33(4):e13719. doi:10.1111/dth.13719

33. Seethapathy H, Zhao S, Chute DF, et al. The Incidence, Causes, and Risk Factors of Acute Kidney Injury in Patients Receiving Immune Checkpoint Inhibitors. Clinical Journal of the American Society of Nephrology. 2019;14(12):1692–1700. doi:10.2215/CJN.00990119

34. Cortazar FB, Kibbelaar ZA, Glezerman IG, et al. Clinical Features and Outcomes of Immune Checkpoint Inhibitor-Associated AKI: A Multicenter Study. Journal of the American Society of Nephrology. 2020;31(2):435–446. doi:10.1681/ASN.2019070676

35. Gupta S, Short SAP, Sise ME, et al. Acute kidney injury in patients treated with immune checkpoint inhibitors. J Immunother Cancer. 2021;9(10):e003467. doi:10.1136/jitc-2021-003467

36. Cortazar FB, Marrone KA, Troxell ML, et al. Clinicopathological features of acute kidney injury associated with immune checkpoint inhibitors. Kidney Int. 2016;90(3):638–647. doi:10.1016/j.kint.2016.04.008

37. Tang SQ, Tang LL, Mao YP, et al. The Pattern of Time to Onset and Resolution of Immune-Related Adverse Events Caused by Immune Checkpoint Inhibitors in Cancer: A Pooled Analysis of 23 Clinical Trials and 8,436 Patients. Cancer Research and Treatment. 2021;53(2):339–354. doi:10.4143/crt.2020.790

38. De Martin E, Michot JM, Papouin B, et al. Characterization of liver injury induced by cancer immunotherapy using immune checkpoint inhibitors. Journal of Hepatology. 2018;68(6):1181–1190. doi:10.1016/j.jhep.2018.01.033

39. Couey MA, Bell RB, Patel AA, et al. Delayed immune-related events (DIRE) after discontinuation of immunotherapy: diagnostic hazard of autoimmunity at a distance. Journal for ImmunoTherapy of Cancer. 2019;7(1):165. doi:10.1186/s40425-019-0645-6

40. Khoja L, Day D, Wei-Wu Chen T, Siu LL, Hansen AR. Tumour- and class-specific patterns of immune-related adverse events of immune checkpoint inhibitors: a systematic review. Ann Oncol. 2017;28(10):2377–2385. doi:10.1093/annonc/mdx286

41. Xu C, Chen YP, Du XJ, et al. Comparative safety of immune checkpoint inhibitors in cancer: systematic review and network meta-analysis. BMJ. 2018;363:k4226. doi:10.1136/bmj.k4226

42. Haanen J, Obeid M, Spain L, et al. Management of toxicities from immunotherapy: ESMO Clinical Practice Guideline for diagnosis, treatment and follow-up. Ann Oncol. 2022;33(12):1217–1238. doi:10.1016/j.annonc.2022.10.001

43. Alatawi YM, Hansen RA. Empirical estimation of under-reporting in the U.S. Food and Drug Administration Adverse Event Reporting System (FAERS). Expert Opin Drug Saf. 2017;16(7):761–767. doi:10.1080/14740338.2017.1323867

44. Neha R, Subeesh V, Beulah E, Gouri N, Maheswari E. Existence of Notoriety Bias in FDA Adverse Event Reporting System Database and Its Impact on Signal Strength. Hosp Pharm. 2021;56(3):152–158. doi:10.1177/0018578719882323

45. Kreimeyer K, Spiker J, Dang O, De S, Ball R, Botsis T. Deduplicating the FDA adverse event reporting system with a novel application of network-based grouping. J Biomed Inform. 2025;165:104824. doi:10.1016/j.jbi.2025.104824

