## Supplementary material for "Immune checkpoint blockade reshapes drug-associated toxicity: a pharmacovigilance atlas of drug-ICI interactions": Online Supplement

**ONLINE SUPPLEMENTAL MATERIAL**

Contents: eMethods; eTables 1-13 (provided in Supplemental_Tables_eTables1-13.xlsx); eFigures 1-6 (provided in eFigure_COMBINED.pdf).

**eMethods. FAERS preprocessing, canonical drug assignment, cancer-associated report ascertainment, phenotype definitions, and statistical analysis**

Preprocessing was performed in three sequential layers before statistical modeling: (1) package-native FAERS acquisition, standardization, and report-level deduplication; (2) provenance-preserving normalization of FAERS drug labels to canonical medicinal exposures; and (3) independent ascertainment of cancer-associated reports from reviewed indication and cancer-specific drug-proxy dictionaries. The deduplicated report universe and reviewed dictionaries were frozen before construction of the analytic exposure and outcome matrices.

### FAERS acquisition, standardization, and deduplication

#### Package-native ingestion and standardization

We constructed the 2016–2025 source dataset using the Bioconductor faers package (version 1.8.0). All 40 quarterly FAERS ASCII releases from 2016Q1 through 2025Q4 were downloaded and parsed with faers::faers_parse(). Quarter-level objects were then combined with faers::faers_combine() before deduplication. Combining the full 10-year series first was intentional so that reports repeated across quarterly releases could be handled within the same deduplication operation rather than only within individual quarters.

The combined FAERS object was standardized with faers::faers_standardize() using MedDRA version 29.0; standardized Standardised MedDRA Query annotations were enabled during this step. The package-native relational structure was retained, including demographic (DEMO), drug (DRUG), reaction (REAC), outcome (OUTC), indication (INDI), therapy (THER), and report-source (RPSR) tables.

#### Report-level deduplication

Report-level deduplication was performed after combination and standardization using faers::faers_dedup(data_standardized, remove_deleted_cases = TRUE). Thus, identification and resolution of duplicate FAERS reports were delegated to the package-native deduplication implementation rather than to a study-specific fuzzy-matching or record-linkage heuristic, and cases identified by the source as deleted were removed. No quarter-specific deduplication was substituted for this multi-year operation.

The resulting deduplicated FAERS universe contained 13,701,106 reports and served as the fixed report-level substrate for all analyses in this study. Downstream preprocessing sometimes collapsed repeated links within a report—for example, repeated appearances of the same canonical ingredient were reduced to a single report–ingredient exposure—but these operations did not create new report identities or constitute an additional report-level deduplication procedure. For storage, character columns were converted to valid UTF-8, nonprinting control characters were removed, and leading or trailing whitespace was normalized without altering the relational report identifiers.

### Age Handling and Imputation

Age at report was analyzed as a continuous variable. For reports with missing age, we performed sequential hot-deck imputation using the impute_shd function in the simputation R package with the VIM backend and a fixed random seed of 111. The donor model included sex, geographic region, calendar year, number of reported drugs, cancer status, ICI exposure, reported death, and the prespecified focal adverse-event phenotypes. These variables were specified in advance rather than selected empirically from the observed data. Report identifiers, therapy dates, individual drug exposures, and other high-dimensional drug features were not used for donor selection.

Imputation was performed only for reports with missing age; all originally observed ages were retained unchanged. The original missing-age indicator was also preserved and included as a covariate in adjusted regression models to account for potential differences between reports with observed and missing age. Imputed ages were required to fall within the prespecified allowable range of 0 to 120 years, and the completed age variable contained no missing values. In the adjusted models, age was represented using a natural spline with 4 degrees of freedom rather than assuming a linear association with the outcome.

Because hot-deck imputation provides a single completed age value rather than multiple-imputation estimates combined using Rubin rules, robustness to age handling was additionally assessed by repeating the primary drug-by-ICI interaction analyses among reports with originally observed age only (eFigure 5; eTable 11).

### Drug normalization and canonical ingredient mapping

Within the deduplicated 2016–2025 FAERS dataset, we developed a de novo, provenance-preserving pipeline to standardize drug exposures in the DRUG table to a common canonical medicinal-exposure ontology. The pipeline began with 52,532,690 original DRUG rows. Each row was assigned a stable parent_row_id corresponding to its physical position in the source table, permitting all subsequent transformations to be propagated deterministically back to the original record. Original FAERS fields, including primaryid, drug_seq, drugname, prod_ai, drug role, and available dose and formulation information, were retained for provenance. For mapping, prod_ai was preferentially used when it contained usable drug information; when prod_ai was missing, blank, or consisted only of clearly non-drug information, drugname was used instead. Both original fields were retained regardless of which field supplied the mapping label. This approach was designed to preserve the original FAERS row universe rather than replacing or deleting source records.

#### Pre-LLM parsing and normalization

Source labels underwent conservative parsing before semantic adjudication. Because individual FAERS entries can contain multiple drugs or ingredient fragments in a single field, selected delimiters were used to create component-level mapping instances while preserving the unsplit parent record. Commas were not treated as universal delimiters because they frequently occur in strengths, product descriptions, manufacturer information, and other legitimate drug-name syntax. Backslashes and semicolons could serve as component delimiters, while forward slashes were split only after protecting recognized non-delimiter uses, including concentrations and ratios, dates and fractions, influenza strain nomenclature, manufacturer notation, factor VIII/von Willebrand factor nomenclature, and other drug-specific constructs. Delimiters occurring within protected bracketed or parenthetical expressions were not blindly split. Square-bracket content could contribute explicit ingredient evidence in defined parsing contexts, whereas parenthetical content was retained primarily as contextual evidence. Parsing decisions and potentially ambiguous transformations were stored as provenance flags, allowing the original source representation to remain recoverable. The resulting component-level table contained 57,780,605 mapping instances linked to the 52,532,690 original DRUG rows.

Component strings then underwent loss-minimizing deterministic normalization. Processing included Unicode normalization, uppercasing, whitespace normalization, normalization of selected symbols and units, and removal of high-confidence nonidentity information such as explicit strengths or concentrations, package quantities, dosing instructions, and clearly administrative route, frequency, or dosage-form annotations. Potentially identity-bearing punctuation, intrinsic numbers, hyphens, parentheses, and protected slashes were otherwise retained rather than aggressively stripped. A separate punctuation-insensitive matching representation was generated to group lexical variants without requiring the displayed clean label itself to discard potentially meaningful information. The actual adjudication input consequently retained some incompletely normalized or malformed labels when aggressive removal could have destroyed medicinal identity; such residual complexity was intentionally deferred to semantic adjudication. Parsing provenance such as FORWARD_SLASH_FISSION, SEMICOLON_FISSION, and foundation review flags remained available to later review.

Clearly unusable component strings were excluded deterministically before model adjudication. These rules were limited to high-specificity classes such as placeholders, pure dates or numbers, punctuation-only or unit-only strings, strength- or concentration-only entries, route-only entries, packaging-only entries, isolated dosing instructions, and strings rendered empty by deterministic cleanup. Exclusion reason codes were retained rather than treating these records as missing data. This rule-based stage excluded 2,670 of the 57,780,605 component-level mapping instances; uncertain or potentially medicinal labels were deliberately retained for semantic adjudication rather than being discarded by aggressive string rules.

#### Construction of adjudication strata

To avoid repeatedly adjudicating the same lexical exposure across tens of millions of records, retained mapping instances were collapsed into 290,834 unique adjudication strata. Strata grouped compatible normalized representations while retaining the underlying component-to-parent crosswalk. A representative cleaned label (ingredient_raw) was selected for each stratum, and alternate observed punctuation or spelling forms were retained as label_variants. Report burden was calculated as the number of distinct primaryid values represented by each stratum rather than the number of component rows, preventing repeated mentions within the same report from inflating frequency.

The model input was intentionally contextual but compact. Each stratum contained an immutable LLM row identifier and adjudication-stratum key, the representative label, alternate label variants, distinct-report count, distribution of reporting countries, the proportion of observations originating from prod_ai versus drugname, source/origin provenance, parsing flags, and available bracket, parenthetical, product-prefix, or related parsing evidence. These were the fields actually supplied to the language model; indication, route, dosage form, application number, and external candidate dictionaries were not included as separate model-input features in this implementation. Related lexical strata were ordered together before adjudication so that spelling, salt, formulation, and related label families tended to occur in neighboring batches.

#### GPT-5.6 Sol canonical adjudication

The 290,834 strata were adjudicated between August 12 and 15, 2026 using OpenAI GPT-5.6 Sol in batches of up to 1,000 strata. A fixed adjudication prompt instructed the model to evaluate every input stratum and return a structured record containing suggested_canonical, adjudication status, mapping type, confidence, country-specific information when relevant, the number of canonical components, an indicator of unintentionally concatenated multiple drugs, an indicator for human review, and a rationale. Permitted adjudication states were ACCEPT, EXCLUDE, COUNTRY_SPLIT, and MANUAL_REVIEW; canonical medicinal names were required to be uppercase, and true multi-active combinations were represented as alphabetized ingredients separated by semicolons.

The prompt specified a common pharmacologic ontology rather than asking the model merely to standardize spelling. Unmistakable generic names, spelling variants, brands, fixed combinations, biologics, biosimilars, insulins, vaccines, vitamins, minerals, supplements, herbals, and broad medicinal exposures were all adjudicated in the same comprehensive pass. Routine salts, counterions, and solvates were generally reduced to the underlying active ingredient when they did not define a clinically meaningful exposure. In contrast, clinically important forms were preserved when collapsing them would conflate distinct exposures; prespecified examples included depot and long-acting formulations, important prodrugs, distinct therapeutic iron and calcium compounds, selected corticosteroid esters, and other clinically meaningful distinctions such as metoprolol tartrate versus metoprolol succinate. Biologically modified products were retained separately when appropriate, whereas nonmeaningful biosimilar suffixes could be normalized to the parent biologic. Vaccines were represented primarily at coherent vaccine-product or vaccine-family levels rather than decomposed into individual antigen or strain fragments. The model was additionally instructed to detect unintentionally concatenated multiple-drug labels without oversplitting salt names, prodrugs, biologic nomenclature, vaccine terminology, or brand-plus-generic duplicates.

Brand mappings were resolved to active ingredient or ingredients when medicinal identity was sufficiently supported. Because identical proprietary names can denote different products in different countries, report-country context was explicitly available during adjudication. A brand with a common medicinal identity across represented markets could receive a global mapping; a brand with materially different active ingredients across markets was assigned COUNTRY_SPLIT rather than creating an artificial combination of the different national formulations. When a label supported a genuine medicinal exposure but not a defensible molecular identity, the ontology permitted a broader clinically interpretable concept, such as an unspecified medication class, vitamin/mineral preparation, laxative, corticosteroid, antihistamine, antacid, or nutritional supplement, rather than requiring either an invented ingredient or automatic exclusion.

#### Post-LLM validation

All model-output files were reassembled using the immutable LLM row identifier and adjudication-stratum key. The expected and observed universes each contained 290,834 unique strata, with no missing strata, unexpected strata, duplicate stratum assignments, or identifier mismatches. Output-file and within-file row provenance were retained.

The combined output then underwent programmatic validation before any human decisions were applied. Checks included permitted adjudication and confidence values, required canonical names for accepted mappings, absence of canonical values for exclusions, uppercase canonical formatting, consistency between combination strings and reported component counts, internal consistency of country-specific decisions, and other schema contradictions. Nonstandard but interpretable model-generated mapping-type labels were standardized to the project vocabulary rather than automatically being treated as substantive mapping failures. Two otherwise correct beta-glucan spelling mappings contained malformed metadata because mapping-type and confidence information had been returned in incorrect fields; these two records were repaired deterministically while preserving their original values as provenance.

#### Targeted human adjudication

Human review was targeted to mappings most likely either to contribute materially to the dataset or to propagate an uncertain model decision. Rather than manually reviewing the entire long tail of rare unresolved labels, we constructed a 934-stratum priority queue. This included 427 MANUAL_REVIEW strata occurring in at least 25 distinct FAERS reports, 410 mappings that GPT-5.6 Sol had classified as ACCEPT but assigned LOW confidence, 95 COUNTRY_SPLIT strata, and the 2 deterministic metadata repairs described above. Thus, 932 strata underwent substantive targeted human review, while 2 additional records were retained in the queue as deterministic repair artifacts. Importantly, review therefore included not only mappings the model could not resolve, but also mappings the model had proposed accepting while expressing low confidence, providing an explicit check against uncertain false-positive canonical assignments. The review strategy was based on the model adjudication and report burden rather than solely on the model's own human_review_required indicator.

For human review, the compact queue was augmented with the same contextual evidence used during GPT adjudication, including alternate label variants, country distribution, prod_ai versus drugname provenance, parsing origin, parsing flags, and other retained evidence context. Reviewers could confirm or revise the proposed ingredient, assign a broader defensible ontology concept, exclude a nonmedicinal or unusable label, or retain country-specific resolution when different markets represented genuinely different exposures. Human review followed the same core ontology used in the model pass, including reduction of routine salts, preservation of clinically meaningful forms, coherent handling of combinations and complex medicinal products, and use of broad unspecified concepts only when the raw label supported that medicinal class. Human decisions superseded the corresponding model decisions, while the original pre-human adjudication, canonical exposure, mapping type, confidence, and rationale were retained for audit.

Completed review files underwent a second set of programmatic integrity checks before being incorporated. Every reviewed record was required to join one-to-one to the existing adjudication table by permanent stratum key, and its LLM row identifier and raw representative label were independently checked for identity. Human ACCEPT decisions required a canonical exposure; EXCLUDE and unresolved decisions could not retain a stale canonical exposure; and retained COUNTRY_SPLIT decisions required a usable country map.

#### Country-dependent drug resolution

The 95 country-review strata were individually adjudicated to determine whether national formulations required different canonical assignments. Where all known formulations could be represented by a narrower common medicinal class, the label was instead assigned a global ontology rollup; where the medicinal identity differed materially by country, a country-specific mapping was retained. The final review artifacts consequently contain both patterns—for example, some market-variable antacid or supplement products were rolled up to a common clinically meaningful exposure, whereas products representing distinct prescription ingredients in different countries retained separate country-specific mappings.

Country maps were normalized to a strict representation using two-character country codes. Legacy full-country names, alternate serialized formats, composite country keys, and spreadsheet-escaped JSON were normalized before application. Noninformative fallback categories such as OTHER, unknown, or unspecified country were not used to infer a formulation from another market.

For mappings retaining a country split, report country was obtained from FAERS DEMO using the prespecified hierarchy of occurrence country, followed by reporter country, followed by the general country field. If duplicate DEMO records for the same primaryid contained conflicting known countries, missing values were ignored, the most frequently occurring known country was selected, and alphabetical order was used to break ties. Country lookup was performed only for reports containing a country-sensitive mapping. A country-specific ingredient was applied only when the report's normalized two-character country code was explicitly represented in the reviewed country map; no formulation was borrowed from another country when the report country was missing or unlisted.

#### Reintegration into original FAERS DRUG rows

Final stratum-level decisions were propagated through the saved 57,780,605-row component crosswalk rather than by rerunning the original parsing procedure. The original DRUG and DEMO source files were checksum-verified before reintegration to ensure that parent_row_id still referred to the same source-row ordering. Each component inherited the finalized stratum adjudication, with deterministic pre-LLM exclusions retained as exclusions and retained country splits first resolved using report country.

For original DRUG rows that had generated multiple mapping components, the component decisions were then recombined conservatively. If one or more components were accepted and no medicinal component remained unresolved, accepted canonical components were split into their constituent semicolon-delimited ingredients, deduplicated, alphabetized, and recombined into a single semicolon-separated exposure string. Excluded nonmedicinal parsing fragments therefore did not invalidate an otherwise interpretable drug entry. Conversely, if any potentially medicinal component remained unresolved, the entire parent row was initially retained as unresolved rather than silently dropping the uncertain component. A parent for which all components were excluded was classified as EXCLUDE. The reconstructed mapping was then attached to the corresponding original DRUG row without deleting or altering the original FAERS source columns.

Following targeted human review and country-specific resolution, the remaining unresolved long tail was treated as a terminal exclusion rather than subjected to further low-yield manual adjudication. These rows retained their pre-terminal unresolved status as provenance but received a blank canonical exposure and final row-level mapping_final_adjudication = "EXCLUDE". Consequently, the final archival DRUG table preserves all 52,532,690 original FAERS DRUG rows, while the authoritative downstream mapping state is binary: accepted rows contain a canonical medicinal exposure, and excluded rows contain no canonical exposure. Final integrity checks required every ACCEPT row to have a canonical exposure, every non-ACCEPT row to have none, and all canonical strings to conform to the uppercase ontology.

#### Construction of drug-exposure variables

Downstream analyses did not treat excluded rows as drug exposures. Only original DRUG rows with mapping_final_adjudication = "ACCEPT" and a nonmissing canonical exposure were admitted to exposure matrices. For canonical mappings containing multiple semicolon-separated active ingredients, the combination was expanded to its constituent canonical ingredients and deduplicated to one observation per report and canonical ingredient before construction of the binary sparse exposure matrix. Thus, a report containing the same canonical ingredient through multiple FAERS DRUG rows contributed a single positive exposure indicator for that ingredient. The same final-adjudication rule was applied to analyses restricted to primary-suspect drugs: primary-suspect rows classified as EXCLUDE did not contribute to the primary-suspect exposure matrix.

The completed pipeline assigned a usable canonical exposure to 51,975,247 of 52,532,690 original DRUG rows (98.94%); 557,443 rows (1.06%) were excluded from ingredient-based analyses. Because the excluded records were retained in the archival mapping table rather than physically deleted, all source rows and mapping decisions remained auditable while downstream analyses were restricted to accepted canonical exposures.

### Cancer-associated report ascertainment

Cancer status was assigned at the FAERS report level and was defined independently of the adverse-event outcomes analyzed in this study. A report was classified as cancer associated if it contained either (1) a reviewed malignant indication or (2) an accepted canonical drug whose presence alone was judged to be highly specific evidence that the patient had cancer. The latter criterion was intentionally stricter than identifying drugs that can be used in oncology. The final binary cancer flag therefore represented evidence that the report arose from a cancer treatment context rather than a broad list of medications with any antineoplastic use.

#### Reviewed cancer-indication dictionary

We first constructed a dictionary of the distinct MedDRA indication codes and Preferred Term labels observed in the FAERS INDI table. An automated high-specificity screen identified explicit malignant language and disease identities that are intrinsically malignant, including solid-organ cancers and recognized hematologic malignancies such as leukemia, lymphoma, myeloma, myelodysplastic syndromes, and myeloproliferative neoplasms. These hematologic entities did not require the literal word “malignant” to qualify. Generic terms such as neoplasm, tumor/tumour, organ-specific neoplasm, adenoma, thymoma, glioma, carcinoid, and other entities with potentially benign or malignant forms were routed to manual review rather than automatically classified as cancer. Terms explicitly indicating benign or uncertain behavior and lexical false positives such as malignant hypertension, malignant hyperthermia, tumor necrosis factor, body mass, and neoplasm prophylaxis were not automatically called cancer.

The complete indication dictionary was manually adjudicated where required, with a final binary cancer_indication_manual decision retained for every exported indication code. The review distinguished a malignant diagnosis from related treatment or context terms; for example, generic chemotherapy, adjuvant therapy, neoadjuvant therapy, and prophylactic chemotherapy were not by themselves treated as malignant indications, whereas specific malignant neoplasms and clinically malignant organ-neoplasm terms were retained. Programmatic checks prohibited blank decisions, contradictory decisions for the same MedDRA code, or omission of any indication code from the reviewed decision universe.

#### Reviewed cancer-specific drug-proxy dictionary

A second, independent route to cancer ascertainment used canonical drug exposures. Candidate proxy drugs were generated by linking INDI rows to the specific FAERS DRUG row using primaryid plus drug_seq and then joining those drug rows to the finalized canonical ingredient mapping. Every canonical ingredient linked to at least one cancer-positive indication was exported for review; evidence included total report burden, cancer-linked and noncancer-linked indication counts, primary-suspect cancer-linked counts, ICI-associated counts, and the most frequent linked malignant indications. A review-priority label was used only to order the adjudication file and did not remove lower-priority ingredients from the candidate universe.

The manual proxy question was deliberately narrow: would observing this ingredient, by itself, provide sufficiently specific evidence that the patient had cancer? Accordingly, drugs with substantial nononcologic use were not accepted merely because they can be part of cancer therapy. Examples of deliberately negative mixed-use proxies included methotrexate, mycophenolate, infliximab, rituximab, cyclophosphamide, and systemic corticosteroids. In contrast, sufficiently cancer-specific antineoplastic agents could be accepted even when the corresponding report lacked a usable malignant indication. All agents in the curated ICI dictionary were required to remain positive cancer proxies, enforcing the prespecified invariant that an ICI-containing report was cancer associated.

Each proxy candidate received a final binary cancer_proxy_manual decision. Canonical ingredient names were normalized before application, and the analysis halted if the same ingredient had conflicting decisions, if a reviewed candidate was missing from the final dictionary, or if any curated ICI had been assigned a negative proxy decision.

#### Final report-level cancer flag and audit

Reviewed positive indication codes were projected to the report level through the binary indication matrix, and reviewed positive cancer-proxy drugs were projected through the all-role canonical drug-exposure matrix. The final cancer indicator was defined as cancer_flag = 1 when either cancer_indication_flag = 1 or cancer_proxy_drug_flag = 1. Drug role was not required for cancer-proxy ascertainment because the purpose was to identify treatment context rather than causal attribution. An indication-only flag was retained separately for audit and sensitivity work.

The final cancer definition identified 2,365,278 of 13,701,106 deduplicated FAERS reports as cancer associated; 256,940 of these contained an ICI in any drug role. Automated integrity checks required the final cancer flag to be a superset of both component flags, required the indication-only flag to reproduce the indication component exactly, and tabulated the incremental number of reports contributed by cancer-specific drug proxies. Because cancer ascertainment used only INDI and canonical DRUG information, focal adverse-event phenotypes were not used to determine cancer status.

### Focal Adverse-Event Phenotype Definitions

Focal adverse-event phenotypes were defined using Medical Dictionary for Regulatory Activities (MedDRA) Preferred Term (PT) codes. A report was considered positive for a phenotype if it contained at least 1 of the corresponding PT codes. The definitions used throughout the interaction and temporal analyses were as follows:

- Stevens-Johnson syndrome/toxic epidermal necrolysis (SJS/TEN): 10042033 (Stevens-Johnson syndrome), 10044223 (Toxic epidermal necrolysis), 10083164 (SJS-TEN overlap), and 10091034 (Atypical Stevens-Johnson syndrome).
- Drug reaction with eosinophilia and systemic symptoms (DRESS): 10073508 (Drug reaction with eosinophilia and systemic symptoms) and 10089003 (AGEP-DRESS overlap).
- Acute generalized exanthematous pustulosis (AGEP): 10048799 (Acute generalised exanthematous pustulosis) and 10089003 (AGEP-DRESS overlap).
- Interstitial nephritis: 10048302 (Tubulointerstitial nephritis).
- Drug-induced liver injury (DILI): 10072268 (Drug-induced liver injury), 10019795 (Hepatitis toxic), and 10071198 (Allergic hepatitis).
- Anaphylaxis: 10002198 (Anaphylactic reaction), 10002199 (Anaphylactic shock), 10002216 (Anaphylactoid reaction), and 10063119 (Anaphylactoid shock).
- Vomiting: 10047700 (Vomiting).

DRESS and AGEP were not treated as mutually exclusive phenotypes. MedDRA PT code 10089003 was intentionally included in both definitions, such that reports meeting this overlapping terminology could contribute to both phenotype groups. Similarly, the broader SJS/TEN and DILI phenotypes were defined as the union of their prespecified component PT codes rather than by text searching at the time of analysis. Anaphylaxis was defined by exact matching to the 4 prespecified PT labels above in the frozen MedDRA reaction dictionary, with the corresponding PT codes resolved programmatically.

### Statistical analysis details

#### Preferred-Term association analysis

To reduce confounding by indication, the primary Preferred Term (PT) association analysis was restricted to reports classified as cancer associated and compared reports containing any ICI in any drug role with cancer-associated reports containing no ICI. PTs were eligible if reported in at least 50 cancer-associated reports overall and at least 10 ICI-containing cancer-associated reports. Reporting odds ratios and 95% CIs were calculated from 2 × 2 contingency tables. Likelihood-ratio P values and Benjamini-Hochberg false discovery rate (FDR) adjustment were computed in log space using 256-bit arbitrary-precision arithmetic to preserve numerical resolution for extremely small probabilities.

#### Higher Level Term network and community detection

Phenotype organization was evaluated within all ICI-containing reports. MedDRA PTs were collapsed to Higher Level Terms (HLTs), and HLTs present in at least 50 ICI-containing reports were retained. Pairwise HLT relationships incorporated co-reporting support, lift, the phi coefficient, normalized pointwise mutual information, and an equal-report-weight sensitivity measure. The primary graph retained HLT pairs with at least 50 co-reports, log2(lift) of at least 1, phi of at least 0.10, normalized pointwise mutual information of at least 0.20, and positive equal-report-weight lift. Contextual nonbiological MedDRA system organ classes were excluded before community analysis. Communities were identified with modularity-based Leiden community detection; Louvain community detection was specified as a software fallback if Leiden was unavailable. Communities were retained as cross-organ when they contained at least 3 HLTs from at least 3 system organ classes and met the prespecified cross-system connectivity criteria. Edge-threshold sensitivity analyses are reported in eTable 9.

#### Primary-suspect drug × ICI interaction models

For each of the 7 focal phenotypes, non-ICI canonical ingredients reported as primary suspect were screened for sufficient support. A drug-phenotype pair was eligible when at least 15 phenotype-positive reports contained the drug and at least 5 phenotype-positive reports contained both the drug and an ICI. The primary model was a report-level logistic regression including primary-suspect drug exposure, any-role ICI exposure, and their interaction. Models adjusted for age using a 4-degree-of-freedom natural spline, the original age-missingness indicator, sex, geographic region, calendar year, number of reported drugs, and cancer status. The exponentiated drug × ICI interaction coefficient was interpreted as the ratio by which the drug-phenotype reporting association differed between ICI-containing and non-ICI reports. Wald P values were calculated with arbitrary-precision arithmetic and Benjamini-Hochberg adjustment was performed separately within each phenotype.

Two complementary robustness analyses were performed. First, the fully adjusted interaction models were refit among reports with originally observed age only. Second, empirical evidence stability was assessed across 5000 repeated random 80% subsamples. The repeated-subsample analysis did not refit the adjusted logistic regression; instead, for each eligible drug-phenotype pair it reconstructed the corresponding phenotype × drug × ICI 2 × 2 × 2 table within each subsample and recomputed the crude interaction contrast. Stability metrics therefore quantify direction and signal persistence under report resampling and are not substitutes for the adjusted primary model.

#### Temporal models

Temporal analyses were restricted to phenotype-positive reports with a complete adverse-event date and at least 1 exact-date primary-suspect treatment start. When more than 1 qualifying primary-suspect start preceded the event, the closest positive start was selected. The primary temporal analysis retained documented onset intervals from 1 through 365 days. Within each phenotype, accelerated failure-time models compared reports with any-role ICI exposure with those without ICI exposure. Weibull, log-normal, and log-logistic distributions were compared using Akaike information criterion, and the distribution with the lowest value was used for that phenotype. Adjusted models included age, original age missingness, sex, geographic region, calendar year, number of reported drugs, and cancer status. Exponentiated coefficients are reported as time ratios; values greater than 1 indicate longer documented onset from the selected primary-suspect treatment start.

A prespecified sensitivity analysis removed the 365-day upper bound while retaining only positive onset intervals. For the checkpoint-pathway analysis, PD-1, PD-L1, CTLA-4, LAG-3, TIGIT, and TIM-3 indicators were entered simultaneously within phenotype-specific accelerated failure-time models, allowing combination regimens to contribute to multiple pathway coefficients. A pathway term was estimable only when at least 20 pathway-positive and 20 pathway-negative observations were available for that phenotype. Benjamini-Hochberg adjustment was applied across all estimable checkpoint-phenotype coefficients.

### Software and reproducibility

#### Analysis environment

Analyses were performed in R version 4.6.1 (2026-06-24 ucrt) in RStudio 2026.08.1+195 “Yellow Yarrow” (Posit Software). Principal R packages and versions were faers 1.8.0, data.table 1.18.4, Matrix 1.7.6, countrycode 1.9.0, stringi 1.8.9, fastglm 0.1.1, Rcpp 1.1.2, Rmpfr 1.1.2, ggplot2 4.0.3, ggrepel 0.9.8, patchwork 1.3.2, igraph 2.3.3, survival 3.8.11, simputation 0.2.9, and VIM 7.0.0. The base splines package was used for natural-spline terms.

### Supplementary table index

eTable 1. Cohort flow counts for main and temporal analyses.

eTable 2. Checkpoint inhibitor agent reports by year, 2016-2025.

eTable 3. Checkpoint target reports by year, 2016-2025.

eTable 4. Checkpoint inhibitor dictionary and target mapping.

eTable 5. Complete cancer-restricted Preferred-Term association results underlying Figure 1.

eTable 6. Cross-organ HLT community membership underlying Figure 2.

eTable 7. Cross-organ community audit metrics underlying Figure 2.

eTable 8. Primary HLT edges retained in Figure 2.

eTable 9. Figure 2 edge-threshold sensitivity analysis.

eTable 10. Complete primary-suspect drug x ICI interaction results underlying Figure 3.

eTable 11. Figure 3 interaction robustness and Figure 5 repeated-subsample stability.

eTable 12. Figure 6 temporal summary, accelerated failure-time results and unrestricted-time sensitivity.

eTable 13. Complete Figure 7 checkpoint-pathway temporality results.

### Supplementary figure legends

**eFigure 1. Cohort and analysis-population flow.** Flow diagram showing the fixed 2016-2025 deduplicated FAERS universe and the analytic populations used for the cancer-restricted Preferred-Term analysis, ICI-containing HLT community analysis, primary-suspect drug x ICI interaction screen, focal-phenotype source population and 1-365-day temporal cohort. PS, primary suspect; TTE, time to event.

**eFigure 2. Checkpoint inhibitor agents and reports by year.** Panel A shows distinct ICI-containing FAERS reports by checkpoint target and calendar year. Panel B shows agent-specific annual report counts on a log10(reports+1) scale. Combination and dual-target agents can contribute to more than one target category.

**eFigure 3. Full-FAERS Preferred-Term association sensitivity analysis.** Role-agnostic ICI versus no-ICI reporting associations across all deduplicated 2016-2025 FAERS reports. The x-axis is the log reporting OR and the y-axis is -log10 Benjamini-Hochberg FDR. This analysis evaluates whether the broad redistribution observed in the cancer-restricted primary analysis persists in the full reporting universe.

**eFigure 4. Community-by-system-organ-class heatmap.** Composition of emergent Higher Level Term communities by primary MedDRA system organ class. Cell labels give the number of HLTs and shading represents the fraction of each community. Asterisks identify the four communities displayed in Figure 2.

**eFigure 5. Complete-case age sensitivity analysis for drug x ICI interactions.** Comparison of interaction coefficients from the primary hot-deck age models with models restricted to reports with originally observed age. Points are colored by the significance and direction of the primary result; the diagonal represents equality.

**eFigure 6. Documented time-to-event distributions and sensitivity to the 365-day analysis window.** Panel A shows empirical distributions for all positive documented intervals from the selected primary-suspect treatment start, without a 365-day upper restriction. Panel B compares adjusted any-ICI time ratios from the primary 1-365-day analysis shown in Figure 6 with the unrestricted positive-interval sensitivity analysis. Both models use the same phenotype-specific accelerated failure-time family.
